# An Interpretable Longitudinal Preeclampsia Risk Prediction Using Machine Learning

**DOI:** 10.1101/2023.08.16.23293946

**Authors:** Braden W Eberhard, Raphael Y Cohen, John Rigoni, David W Bates, Kathryn J Gray, Vesela P Kovacheva

**Author notes:** **CORRESPONDING AUTHOR:** Vesela P. Kovacheva MD PhD, Department of Anesthesiology, Perioperative and Pain Medicine, Brigham and Women’s Hospital, Harvard Medical School, 75 Francis St, L1, Boston, MA 02115. Senior Co-Authors (Kathryn J Gray, Vesela P Kovacheva).

## Abstract

**Background:** Preeclampsia is a pregnancy-specific disease characterized by new onset hypertension after 20 weeks of gestation that affects 2-8% of all pregnancies and contributes to up to 26% of maternal deaths. Despite extensive clinical research, current predictive tools fail to identify up to 66% of patients who will develop preeclampsia. We sought to develop a tool to longitudinally predict preeclampsia risk.

**Methods:** In this retrospective model development and validation study, we examined a large cohort of patients who delivered at six community and two tertiary care hospitals in the New England region between 02/2015 and 06/2023. We used sociodemographic, clinical diagnoses, family history, laboratory, and vital signs data. We developed eight datasets at 14, 20, 24, 28, 32, 36, 39 weeks gestation and at the hospital admission for delivery. We created linear regression, random forest, xgboost, and deep neural networks to develop multiple models and compared their performance. We used Shapley values to investigate the global and local explainability of the models and the relationships between the predictive variables.

**Findings:** Our study population (N=120,752) had an incidence of preeclampsia of 5.7% (N=6,920). The performance of the models as measured using the area under the curve, AUC, was in the range 0.73-0.91, which was externally validated. The relationships between some of the variables were complex and non-linear; in addition, the relative significance of the predictors varied over the pregnancy. Compared to the current standard of care for preeclampsia risk stratification in the first trimester, our model would allow 48.6% more at-risk patients to be identified.

**Interpretation:** Our novel preeclampsia prediction tool would allow clinicians to identify patients at risk early and provide personalized predictions, as well as longitudinal predictions throughout pregnancy.

**Funding:** National Institutes of Health, Anesthesia Patient Safety Foundation.

**RESEARCH IN CONTEXT:** *Evidence before this study:* Current tools for the prediction of preeclampsia are lacking as they fail to identify up to 66% of the patients who develop preeclampsia. We searched PubMed, MEDLINE, and the Web of Science from database inception to May 1, 2023, using the keywords “deep learning”, “machine learning”, “preeclampsia”, “artificial intelligence”, “pregnancy complications”, and “predictive models”. We identified 13 studies that employed machine learning to develop prediction models for preeclampsia risk based on clinical variables. Among these studies, six included biomarkers such as serum placental growth factor, pregnancy-associated plasma protein A, and uterine artery pulsatility index, which are not routinely available in our clinical practice; two studies were in diverse cohorts of more than 100 000 patients, and two studies developed longitudinal predictions using medical records data. However, most studies have limited depth, concerns about data leakage, overfitting, or lack of generalizability.

*Added value of this study:* We developed a comprehensive longitudinal predictive tool based on routine clinical data that can be used throughout pregnancy to predict the risk of preeclampsia. We tested multiple types of predictive models, including machine learning and deep learning models, and demonstrated high predictive power. We investigated the changes over different time points of individual and group variables and found previously known and novel relationships between variables such as red blood cell count and preeclampsia risk.

*Implications of all the available evidence:* Longitudinal prediction of preeclampsia using machine learning can be achieved with high performance. Implementation of an accurate predictive tool within the electronic health records can aid clinical care and identify patients at heightened risk who would benefit from aspirin prophylaxis, increased surveillance, early diagnosis, and escalation in care. These results highlight the potential of using artificial intelligence in clinical decision support, with the ultimate goal of reducing iatrogenic preterm birth and improving perinatal care.

## INTRODUCTION

Preeclampsia is a pregnancy-specific disorder characterized by new onset hypertension and proteinuria after 20 weeks gestation^1^ that complicates 2-8% of pregnancies.^2,3^ In addition to elevated systolic (>140 mmHg) or diastolic (>90 mmHg) blood pressure, patients can develop progressive end-organ damage characterized by proteinuria, elevated liver enzymes, pulmonary edema, seizures, and death. The only definitive treatment is delivery, and as a result, preeclampsia is the leading cause of iatrogenic preterm birth with associated neonatal morbidity and mortality.

Despite extensive research, the upstream causative factors for preeclampsia remain unknown. It is hypothesized that abnormal placental development may be a large contributor.^4^ Current clinical practice is focused on identifying patients at risk based on established clinical criteria, which include demographic characteristics and medical history.^1^ Maternal risk can be mitigated by close surveillance using frequent home blood pressure monitoring and prophylaxis with low-dose aspirin based on the presence of at least one high-risk or two moderate-risk factors.^5^ However, more than half of pregnancies, in particular those in nulliparous individuals, end with preeclampsia in the absence of risk factors.^6,7^ In addition, little is known about the trajectory of risk and the rate of change in risk across pregnancy.

As tools to predict preeclampsia are lacking, multiple predictive models have been developed. Most models utilize clinical risk factors combined with biomarkers, which may not be available in routine care.^6,8^ The majority of these models are also derived from well-curated datasets using statistical methods. ^6–9^ Recently, a few accurate machine learning models have been developed;^9^ however, the full potential of electronic health record (EHR) data has not been realized. Models developed to date have shown promise using small, constrained data sets but have been lacking in predictive power, accessibility, and generalizability. Machine learning models based on the entire EHR have outperformed the rule-based algorithm that relies on established clinical criteria.^10,11^

The modern EHR stores a wealth of patient data and is in use across most health care institutions in the US. In large healthcare systems with multiple hospitals and outpatient offices, using uniform EHR software allows for the detailed aggregation of patient data collected at different encounters, sometimes across different hospitals or outpatient offices, longitudinally. Using the readily available rich data source of the EHR and machine learning to integrate data with complex relationships over time, we sought to develop accurate predictions of the risk for preeclampsia throughout pregnancy. We created a large patient cohort using longitudinal data from six hospitals from a large healthcare system in the Northeastern US. We developed a system of multiple machine-learning models throughout pregnancy and investigated their performance in preeclampsia prediction. Implementing this type of multi-model system to automatically operate within the EHR by ingesting data as it becomes available and generating predictions longitudinally during pregnancy would allow physicians to timely identify, triage, and modify surveillance and levels of care for the highest-risk patients.

## METHODS

### Study population

This study was approved by the Mass General Brigham Institutional Review Board, protocol # 2020P002859, with a waiver of patient consent. We leveraged EHR data from eight hospitals in the New England region with deliveries between February 2015 (when our institution implemented single-vendor EHR across all outpatient offices and inpatient sites) and June 2023. Deliveries within the Mass General Brigham system were selected based on documentation of pregnancy greater than 20 weeks gestation and associated billing codes for cesarean or vaginal delivery.^12^ The final dataset included data from patients for whom longitudinal data starting from the first prenatal visit before 14 weeks of gestation was available, and we analyzed each pregnancy independently.

### Data processing

All data were ingested and processed using our machine learning platform, Medical Record Longitudinal Information AI System (MERLIN),^13^ which extracts, transforms, and harmonizes data from the EHR. We censored our datasets at 14, 20, 24, 28, 32, 36, 39 weeks of gestation, and before the hospital admission for delivery. We selected these time points based on the timing of routinely scheduled outpatient visits when new laboratory and vital signs data are obtained. For each of the eight datasets, we verified by inspecting the time stamps that only data obtained up to the selected time point were included to minimize the risk of data leakage. Regardless of the time point, all data on or after the date of delivery and date of diagnosis (if relevant) were also excluded.

### Model Features and Preeclampsia Phenotype Definition

All demographic, family history, medical history, laboratory studies, medications, insurance, vital signs, and procedural information data were extracted. The demographic features included maternal age, self-reported race and ethnicity, substance use, prenatal care, and insurance information. Features missing over 95% of entries were removed. We selected established risk factors associated with preeclampsia risk in published studies and guidelines. ^7,9–11,14,15^ As International Classification of Diseases (ICD) codes have well-documented limitations, including overall low accuracy in identifying preeclampsia,^4^ we developed a combination of laboratory values, blood pressure measurements, and ICD codes based on the established guidelines^14^ to define the preeclampsia phenotype in our datasets. We validated this approach using 400 records of patients with ICD codes for preeclampsia, which were reviewed by clinical experts (V.K., K.G.); of those, our algorithm correctly identified 94.0% of the cases.

### Model Development

As data were available from eight hospitals, we developed all models using data from six hospitals, two tertiary and four community hospitals (Cohort 1) and reserved the data from two community hospitals (Cohort 2) for external validation; the latter were chosen because they contain a representative sample of the entire population both in size and amount of data available. Additional information about external validation is provided in appendix (p 2).

The remaining cases were again split into training (80%) and testing sets (20%) based on patient identifiers to ensure that a single patient would not have pregnancies in different sets, which could artificially enhance performance metrics. A five-fold cross-validation was performed over the training set to determine hyperparameters. The best combination was then used to calculate testing metrics on the testing set.

We developed the following models, which have been used in prior research.^9–11,16–19^: xgboost, deep neural networks (appendix p 2), elastic net, random forest, and linear regression. Using Optuna’s TPE sampler,^20^ we ran ten iterations over the hyperparameter space to determine the best-fitting combination. These parameters were then used to train the model on the entire training dataset and tested on the testing and external validation sets. We investigated bias, including racial bias, and followed established strategies for mitigation.^21^

We employed Shapley values,^22^ a game theoretic approach to explain the output of any machine learning model. It connects optimal credit allocation with local explanations using the classic Shapley values from game theory and their related extensions. This method allows for both local (for individual patients) and global (for the model) interpretations. We used the SHAP Python package 0.41.0.

### Statistical Analyses and Definitions

For the descriptive analyses, we used all available EHR data from before conception to up to 6 weeks postpartum. Variables were expressed as median with interquartile range (IQR). Significance was determined using the Student’s t-test and one-way ANOVA for parametric variables, the Kruskal-Wallis rank sum test for non-parametric variables, and Fisher’s exact or Chi-squared test for categorical variables. A P-value of less than 0.05 was considered significant.

## RESULTS

### Population characteristics and outcomes

We identified 122,147 deliveries between February 2015-June 2023 in our healthcare system. Of those, 120,752 met the additional selection criteria of having at least one visit with recorded information before 14 weeks of pregnancy and were subsequently used in our analysis. All eight hospitals provided data for this study (appendix p 3) and 62.7% of pregnancies (N=75,740) were in patients receiving care at a tertiary care hospital.

The overall incidence of preeclampsia was 5.7%, N=6,920 (Table 1). Compared to normotensive patients, those with a diagnosis of preeclampsia included a significantly higher proportion of individuals with self-reported Black race (17% vs. 9%), and self-reported Hispanic ethnicity (19% vs 15%), respectively. In addition, patients with preeclampsia were more likely to have a family history of hypertension, higher maximal systolic and diastolic blood pressures, and higher weight gain during pregnancy, compared with patients who did not develop preeclampsia, P<0.01. The remaining characteristics of both groups are summarized in Table 1.

**Table 1.** Characteristics of study patients (total N=120,752)

|  | **Preeclampsia**<br>**(n=6,920)** | **No preeclampsia**<br>**(n=113,832)** | **P-value** |
| --- | --- | --- | --- |
| Maternal age at delivery, y | 32.9 (29.5 - 36.6) | 32.9 (29.9 - 35.8) | 0.17 |
| Self-reported race, White | 4240 (61.3) | 77290 (67.9) | < 0.01 |
| Self-reported race, Black | 1172 (16.9) | 10492 (9.2) | < 0.01 |
| Self-reported race, other | 1553 (22.4) | 26626 (23.4) | 0.11 |
| Self-reported ethnicity, Hispanic | 1289 (18.6) | 17096 (15.0) | < 0.01 |
| Self-reported ethnicity, non-Hispanic | 5631 (81.4) | 96736 (85.0) | < 0.01 |
| Tertiary hospital | 4712 (68.1) | 71028 (62.4) | < 0.01 |
| Community hospital | 2208 (31.9) | 42804 (37.6) | < 0.01 |
| Gravidity | 2.0 (1.0 - 3.0) | 2.0 (1.0 - 3.0) | 0.11 |
| Parity | 1.0 (0.0 - 1.0) | 1.0 (1.0 - 1.0) | < 0.01 |
| Nulliparous | 3207 (46.34%) | 41882 (36.79%) | < 0.01 |
| In vitro fertilization | 414 (5.98%) | 4299 (3.78%) | < 0.01 |
| Multiple gestation | 490 (7.08%) | 2961 (2.6%) | < 0.01 |
| Gestational age at delivery, weeks | 37.7 (36.3 - 39.3) | 39.3 (38.6 - 40.1) | < 0.01 |
| BMI at delivery, kg/m<sup>2</sup> | 33.3 (29.2 - 38.4) | 29.9 (26.9 - 33.8) | < 0.01 |
| Weight at delivery, kg | 87.54 (76.66 - 102.06) | 79.83 (71.21 - 90.72) | < 0.01 |
| Maximal SBP during pregnancy, mmHg | 150 (140 - 160) | 129 (120 - 138) | < 0.01 |
| Maximal DBP during pregnancy, mmHg | 93 (86 - 99) | 80 (74 - 86) | < 0.01 |
| Family history of chronic hypertension | 3075 (44.4) | 41277 (36.3) | < 0.01 |
| Family history of preeclampsia | 142 (2.2) | 2162 (1.9) | 0.14 |
| Past history of chronic hypertension | 1896 (27.4) | 5854 (5.1) | < 0.01 |
| Past history of gestational hypertension | 511 (7.4) | 3528 (3.1) | < 0.01 |
| Past history of preeclampsia | 530 (7.7) | 2205 (1.9) | < 0.01 |
| Past history of preterm delivery<br>(<37 weeks) | 2217 (32.0) | 7747 (5.6) | < 0.01 |
| Antihypertensive medications throughout<br>pregnancy and 6 weeks postpartum | 4767 (68.9) | 6355 (5.6) | < 0.01 |
| Headaches during pregnancy and 6<br>weeks postpartum | 1301 (18.8%) | 12949 (11.38%) | < 0.01 |
| Gestational diabetes | 1247 (18.02%) | 13888 (12.2%) | < 0.01 |
| Proteinuria | 4286 (61.9) | 886 (0.78) | < 0.01 |
| Maximal uric acid during pregnancy | 5.6 (4.7 - 6.6) | 4.6 (3.9 - 5.5) | < 0.01 |
| SGA or IUGR | 579 (8.4) | 9763 (8.6) | 0.56 |

Median (IQR) for continuous variables; n (%) need to convert to percent) for categorical variables; p-values for continuous variables based on Kruskal-Wallis rank sum test; for categorical variables based on Fisher's exact or Chi-squared test.
Abbreviations: SBP, systolic blood pressure; DBP, diastolic blood pressure, BMI, body mass index, SGA, small for gestational age, IUGR, intrauterine growth restriction.

### Longitudinal development of multiple predictive models

To develop predictive models of the preeclampsia risk over time, eight datasets were created at multiple time points in pregnancy from two tertiary and four community hospitals (Cohort 1); the patients from two community hospitals were reserved for external validation (Cohort 2) and those data were excluded from model training (Fig. 1A). The characteristics of both cohorts and all datasets are summarized in the appendix (pp 4-23). For all datasets that were used for model development, data were only included if recorded before the respective cut-off gestational age or before the diagnosis of preeclampsia. The data from patients without preeclampsia were censored at the respective timepoint in pregnancy or before delivery.

**Fig. 1.**
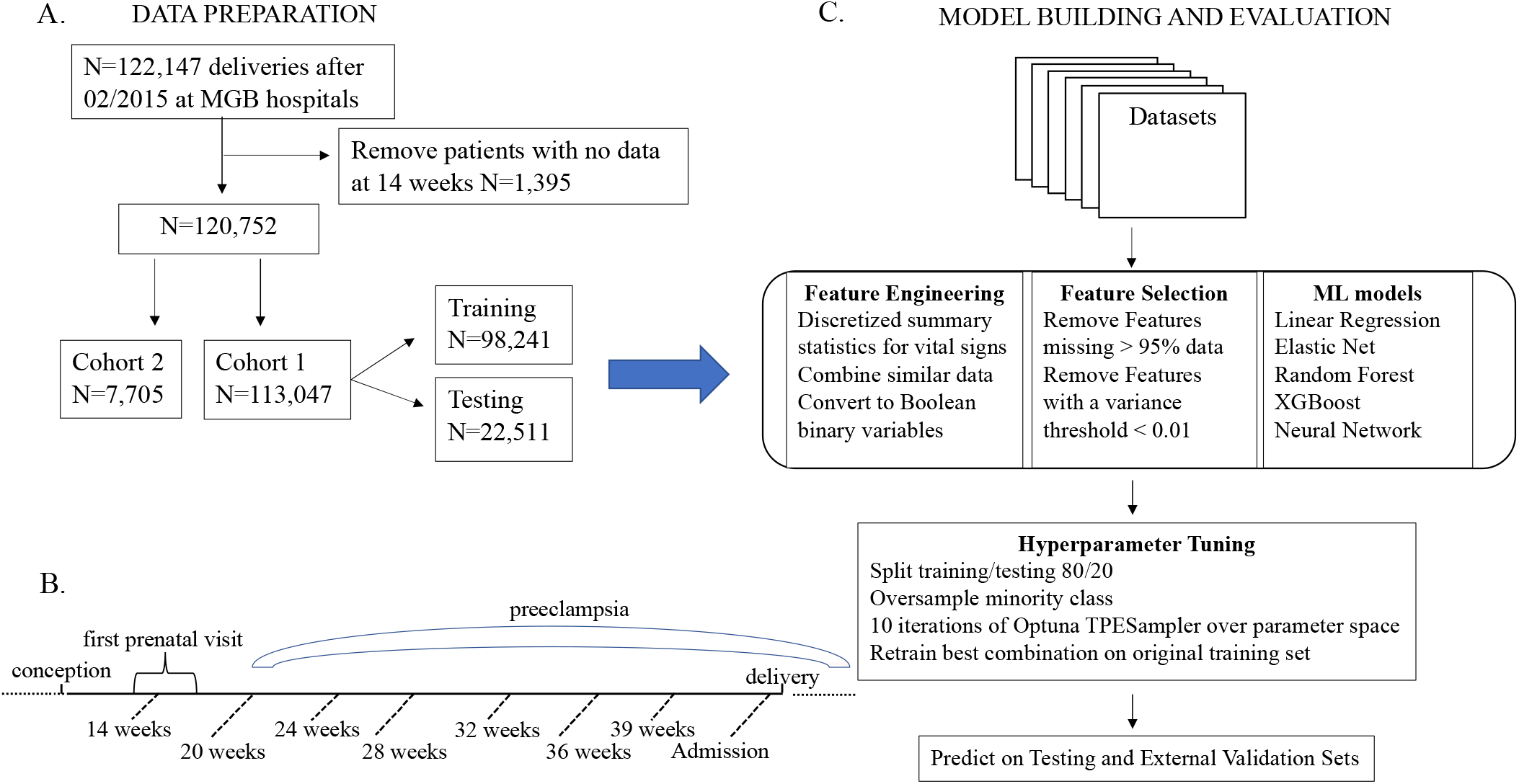
Overview of the data and model development. A. After excluding patients with no data at 14 weeks, the final study patient cohort included 120,752 deliveries; of those, Cohort 1 was divided into training and testing groups used to develop the models, while Cohort 2 was reserved for external validation and did not participate in model development. B. We developed datasets at 14, 20, 24, 28, 32, 36, 39 weeks gestation, and on admission for delivery based on the timing of the routinely scheduled outpatient visits. C. After selecting relevant to the preeclampsia risk features, we developed linear regression, elastic net, random forest, xgboost, and deep neural network models to predict the risk of preeclampsia; the performance of those models was evaluated in the test datasets from Cohort 1 and external validation datasets from Cohort 2.

We developed five types of models: xgboost, deep neural networks, elastic net, random forest, and linear regression at each of the eight different time points: 14, 20, 24, 28, 32, 36, 39 gestational weeks, and on admission (Fig 1B and 1C). The earliest timepoint for which we developed each of the models was 14 weeks, as most patients had their first prenatal visit at that time, and commonly, new vital signs, detailed history, and baseline laboratory tests were obtained. We used the area under the curve (AUC), a widely used metric in machine learning that balances sensitivity and specificity, to evaluate the predictive power of the models.

We analyzed the AUC of each model using the test dataset, which included 20% of the patients in Cohort 1, whose data was not used in the process of model development (Fig. 2A). Overall, the models had AUC 0.73-0.76 at 14 gestational weeks. With the advancement of the gestational age, more data is collected from every patient visit, and the performance of each model type increased; the highest AUC was on admission, range 0.88-0.91. The detailed performance metrics are listed in the appendix (p 24). We externally validated the performance using data from two community hospitals (Fig. 2B and appendix pp 25-26). We also investigated racial bias and optimized the performance of the models (appendix pp 27-28).

**Fig. 2.**
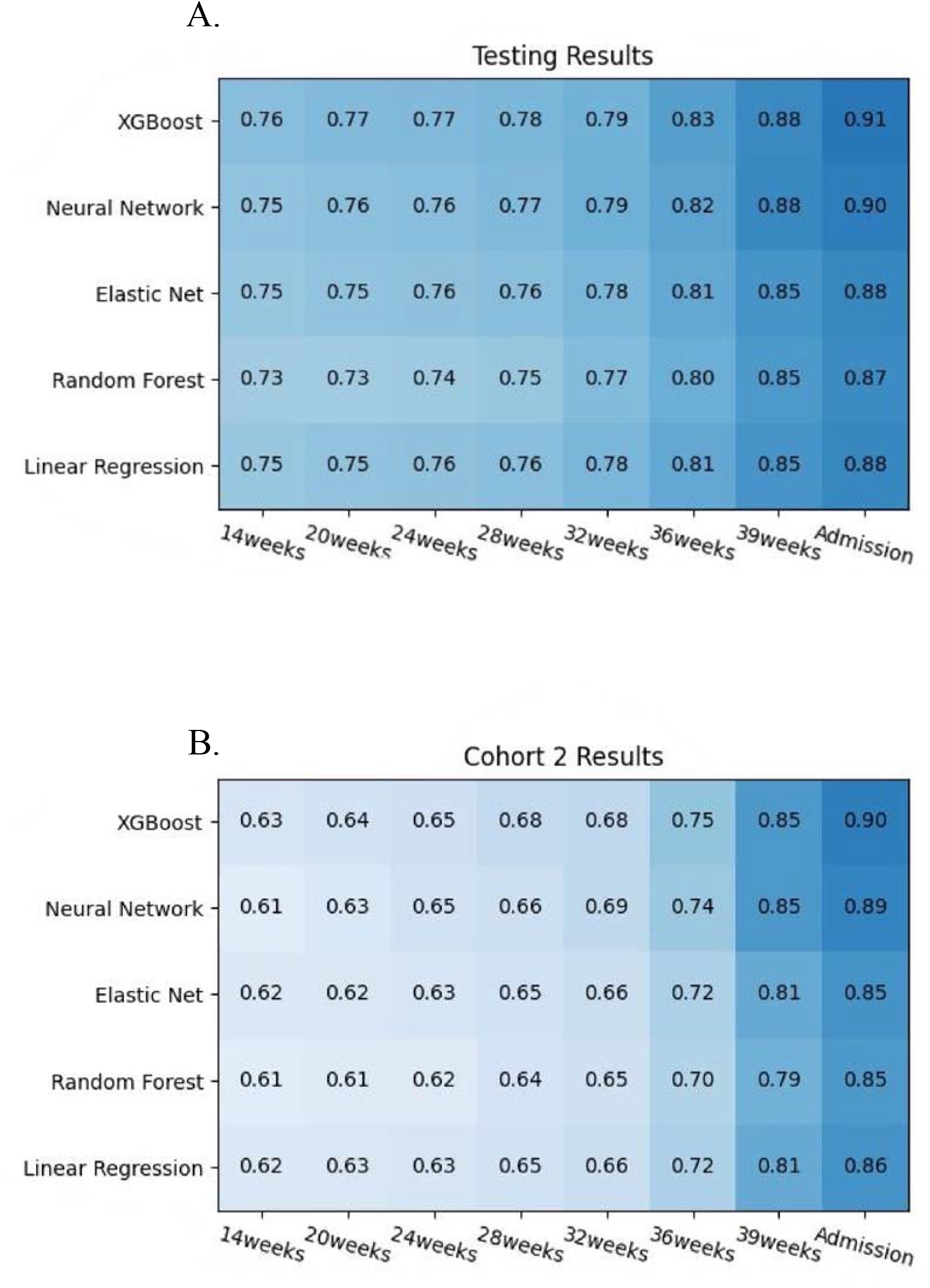
Performance as defined by the area under the curve (AUC) of the xgboost, deep neural network, elastic net, random forest, and linear regression models to predict preeclampsia risk A. Performance on the test dataset from Cohort 1 B. Performance on the external Cohort 2.

### Time course and relationships of the most predictive variables

Using Shapley additive explanations method, we evaluated the most predictive variables in the xgboost model, over the eight time points (Fig 3A). In early pregnancy, at 14 weeks, the most predictive features were chronic hypertension and long interpregnancy interval. With advancing gestational age, the highest contributing features become systolic and diastolic blood pressure. Similarly, in early pregnancy, the category that contributed the most to the model predictions was medical history; with advancing gestational age, the vital signs and laboratory results became more contributory (Fig. 3B). This approach can be used to highlight individual patient predictions and the associated factors (appendix pp 29-30).

**Fig. 3.**
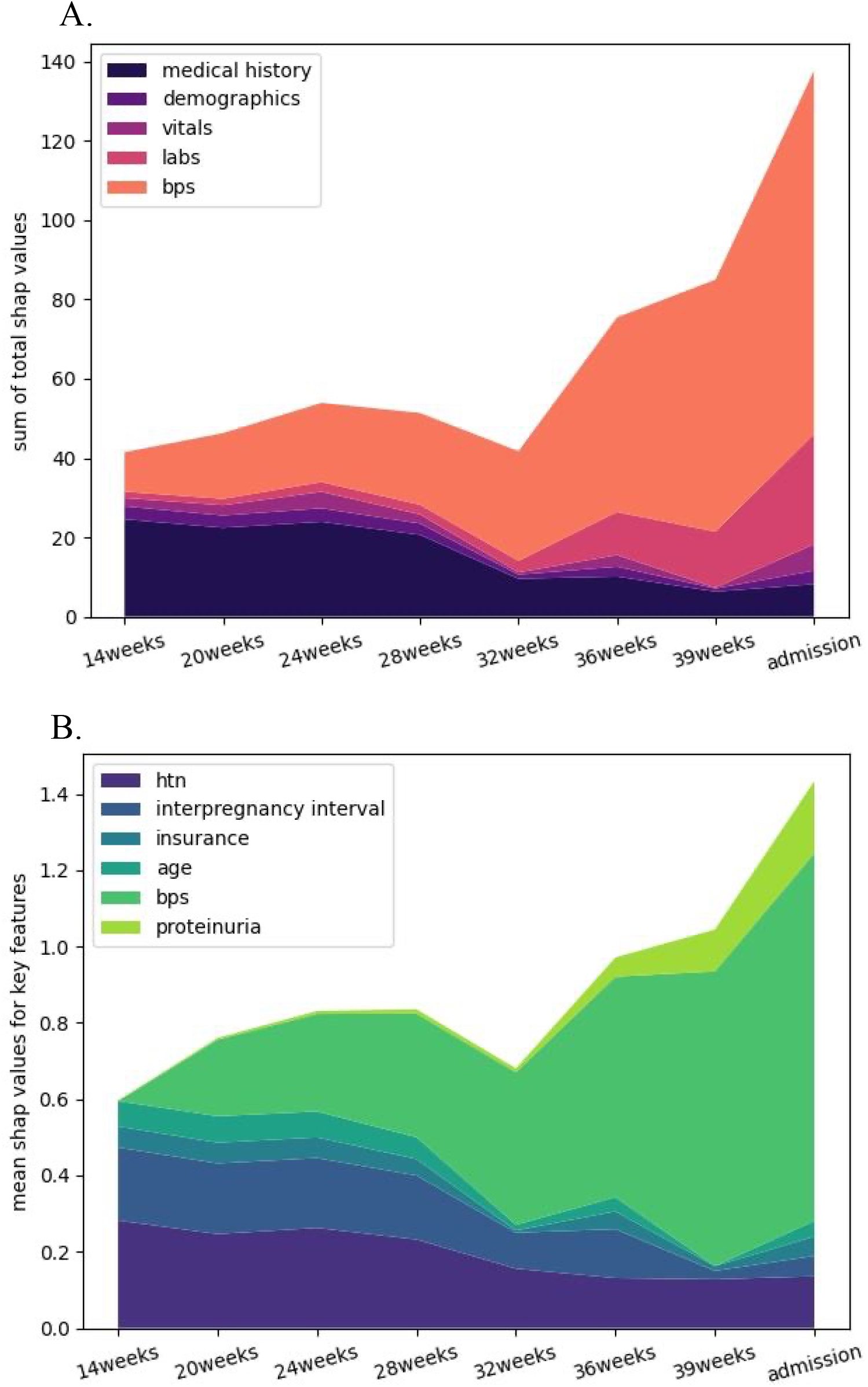
Change in relative contribution for model prediction over time A. Contribution of several feature groups B. The most predictive variables over every timepoint. Labs, laboratory results; bps, systolic and diastolic blood pressure measurements; htn, chronic and gestational hypertension.

In addition to exploring the contribution of individual features to the model’s prediction, Shapley values can highlight the interactions between different variables (appendix p 31). For example, the presence of proteinuria is unlikely to be associated with a diagnosis of preeclampsia if the maximal systolic blood pressure during pregnancy is <140 mmHg; however, above that threshold, proteinuria becomes highly predictive. The relationships between some variables follow non-linear patterns; for example, age is a risk factor if the patient is <20 years or >35 years old, especially if associated with higher parity or higher gravidity. A similar J-shaped relationship is also present between age and short or long interpregnancy intervals. Interestingly, higher SBP is likely to be associated with a higher risk of preeclampsia at age <35 years old. A higher red blood cell count during the second trimester is associated with a higher risk for preeclampsia.

### Comparison to the current standard of care

Although there is no universally accepted model for the prediction of preeclampsia, the ACOG risk stratification criteria are widely accepted in clinical practice^1^. The preeclampsia xgboost model outperformed a model created using the ACOG risk factors based on AUC across all time points (Fig 4A). Moreover, as ACOG guidelines recommend aspirin prophylaxis for all patients at high risk for preeclampsia before 16 weeks, we evaluated how many additional patients would have benefitted from aspirin who would not have been identified by ACOG risk criteria. We compared the performance of our xgboost model with ACOG risk criteria in the test dataset. Using our xgboost model predictions at 14 weeks on the test dataset, 25.0% (N=5624) individuals would be eligible for aspirin prophylaxis, while only 14.6% (N=3295) would be eligible using ACOG criteria. Using the novel preeclampsia model, an additional 2,329 individuals would have been eligible. If aspirin prophylaxis prevents 62% of early-onset preeclampsia in high-risk patients,^23^ an additional 28 cases (total of 66 cases) of early-onset preeclampsia per 10,000 pregnant individuals would have the potential to be prevented with updated risk prediction.

**Fig 4.**
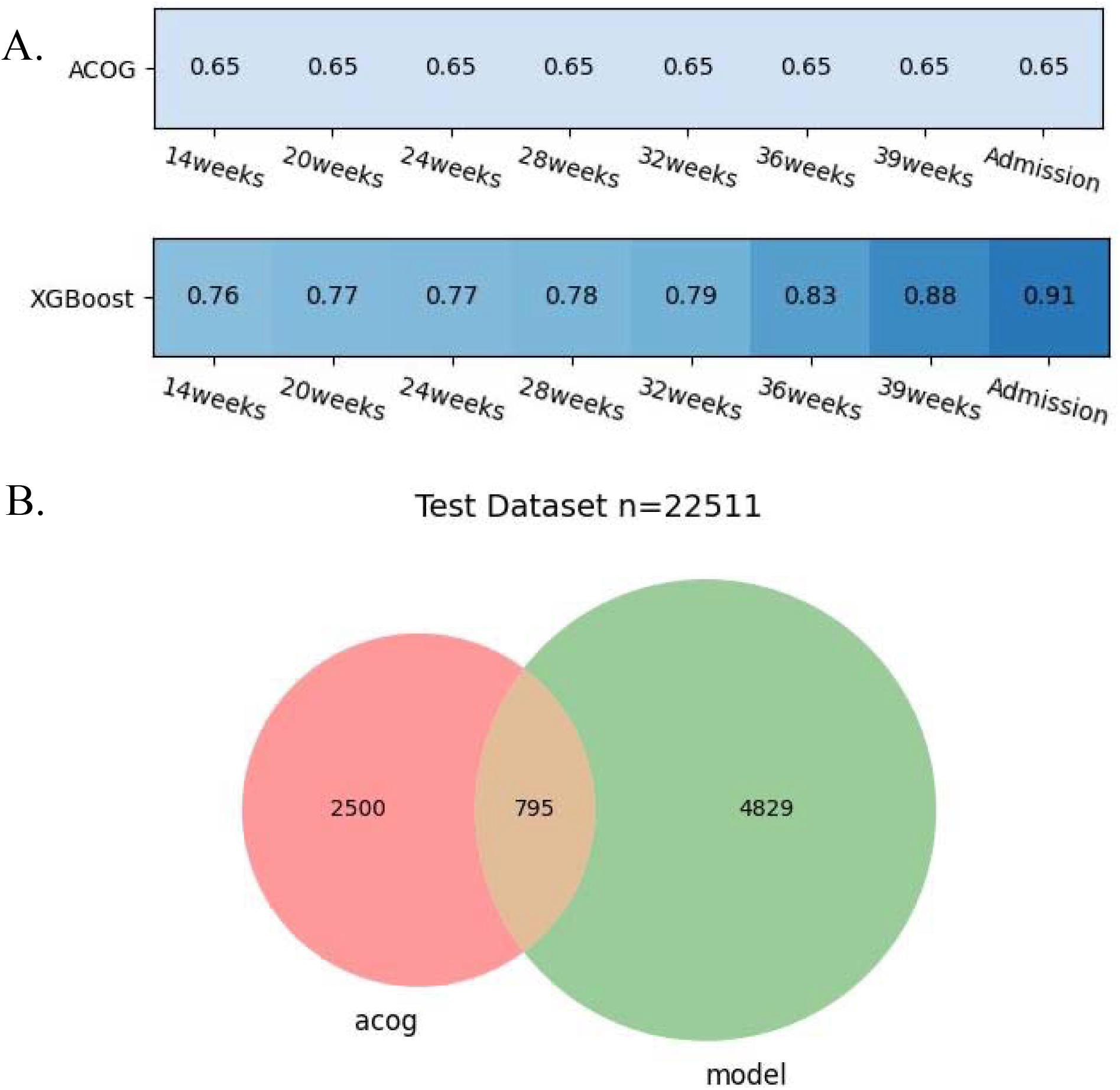
Comparison between the xgboost model and the current standard of care as determined by ACOG guidelines. A. Comparison between AUC of the xgboost model and the ACOG criteria over all time timepoints. B. Venn diagram of all individuals at risk for preeclampsia in the test dataset (N=22,511); of those, individuals at risk predicted using the ACOG criteria (acog) are in pink, and the individuals at risk predicted using the xgboost model (model) at 14 weeks are in green.

## DISCUSSION

We developed an accurate artificial intelligence (AI) tool consisting of eight longitudinal models for preeclampsia prediction using machine learning. With the advancement of pregnancy and the increase in the gestational weeks, the models get progressively more accurate, from area under the curve (AUC) 0.76 to 0.91; the highest predictive value, AUC 0.91, was achieved before delivery. The most predictive features were systolic and diastolic blood pressure and interpregnancy interval. As gestational age advanced, the relative significance of systolic and diastolic blood pressure compared to other predictors increased significantly. Compared with the current standard of care, our models have higher predictive power and would allow earlier and more precise identification of patients at risk for preeclampsia.

The importance of screening for preeclampsia throughout pregnancy has been emphasized,^1^ and to date, multiple predictive models have been developed.^9–11,16–19^ Most studies include clinical data as well as biomarkers, such as serum placental growth factor,^8,19^ pregnancy-associated plasma protein A,^8,19^ and uterine artery pulsatility index.^6,8,19^ However, these tests are not routinely performed in all clinical settings, and thus, the models that utilize these data can be applied only in limited cases. We aimed to develop an accurate tool that can be applied throughout pregnancy and utilizes EHR data routinely collected during prenatal visits. In this way, our AI system has the advantage of being developed using real-world clinical data and can be reproduced in any setting that utilizes EHR.

We achieved high predictive power using our models, which are comparable to or better than other studies using EHR.^6,10,11,18^ We developed multiple models and demonstrated that preeclampsia can be predicted with high performance. The model with highest AUC was xgboost; this type of model has a low risk of overfitting, high interpretability, and performs well in clinical structured data.^24^ In addition, this type of model was highly performant in prior studies predicting preeclampsia.^10,11^ However, these prior studies developed models from all available EHR data, which may lead to overfitting, ^10,18^ did not investigate multiple machine learning models, ^10,11^ and the datasets have high missingness.^11^ In addition, some prior results may display data leakage, for example, including oxytocin, an intra- or post-partum medication in an early antepartum model. We overcome the limitations of prior work by including only those variables which are clinically relevant for the diagnosis of preeclampsia in our models; we anticipate that this approach will improve cross-institutional reproducibility. The external validation and bias investigation are discussed in detail in the Supplemental Discussion (appendix p 32).

The current study is the largest investigation to date based on the cohort size, time points, depth of predictor variables, and types of machine learning models. We demonstrate the strong effect on preeclampsia risk of known predictors, such as a past medical history of preeclampsia, diabetes, and chronic kidney disease.^7,11,16^ In addition, we also incorporated time-series variables in our models. This approach improves the accuracy of predictions,^9,10^ and we replicate prior findings of heightened preeclampsia risk with excessive weight gain^9,25,26^ and steep patterns of blood pressure increase^9,10,27^ during pregnancy. As evidence for the relative role of individual risk factors and interaction between those risk factors is lacking,^26^ our study adds substantial results from clinical practice to inform future predictive tools and guidelines.

The advantage of using Shapley values as an explainable AI approach is that we demonstrate the relative significance of individual factors and the temporal change as the gestational age advances. Not surprisingly, the most important features in early pregnancy are chronic hypertension and interpregnancy interval, which are part of the medical history. Later in pregnancy, the trajectory of the systolic and diastolic blood pressure becomes more important, which potentially may present a subclinical presentation of developing disease. While individual and groups of risk factors have been well investigated,^26^ only a few prior studies have analyzed all patient EHR data during pregnancy in its entirety.^9,10^ Our results offer a more granular look at the individual variables and demonstrate a higher significance of the patient history in early pregnancy, compared to other studies that rank higher vital signs and laboratory results.^10^ These differences likely represent variable institutional practices for early pregnancy screening and highlight the need for future large, multi-center studies.

Preeclampsia risk factors can interact in a complex, non-linear fashion. For example, chronic hypertension is associated with early-onset preeclampsia in patients with weight < 69 kg, no diabetes, and no family history of preeclampsia.^7^ When analyzing the relationships between variables in our models, we demonstrate previously known, as well as new, interactions. The risk of preeclampsia is higher in the extremes of age,^28^ and also in pregnancies with short and long interpregnancy intervals.^29^ The connection with red blood cell count is novel and may be related to a higher hemoconcentration.^30^ As preeclampsia is a heterogeneous disorder, further exploring the interaction between variables has the potential to identify sub-phenotypes of preeclampsia that may benefit from a more personalized approach to prophylaxis and treatment.^31^

Our ability to utilize the Shapley method on an individual level highlights the clinical value of these predictive models to personalize patient management. Identifying the risk factors will allow visualization and potential modification of those factors, which can change the course of the disease and decrease risk. If implemented longitudinally during the pregnancy, for example, in the form of a dashboard, these individualized predictors would allow earlier detection and amelioration in high-risk patients or adjustment of surveillance in low-risk individuals. While individual Shapley values have inherent limitations, this approach can be used to evaluate bias and overall performance.^32^ Across the population, this strategy would allow efficiency and cost savings while maintaining high standards of care.

There is no definitive therapy for preeclampsia (other than delivery); therefore, efforts are focused on prevention and early detection. Multiple studies have shown that aspirin prophylaxis started in early pregnancy in patients at high risk can decrease the incidence of early-onset preeclampsia, which is associated with the highest incidence of maternal and neonatal morbidity.^23,33^ As high-risk patients benefit from aspirin prophylaxis, accurate risk stratification is paramount. The current ACOG guidelines recommend prophylaxis and increased surveillance of high-risk patients;^1^ however, multiple studies have demonstrated that the predictive power of the current standard of care using established risk factors is limited. ^10,11,26^ Therefore, implementing our AI tool in clinical practice may offer substantial benefits for patient health.

Our study has several limitations. In some cases, the diagnosis of preeclampsia may be challenging to establish as there may be no clear tests that distinguish between gestational hypertension, chronic hypertension, and superimposed preeclampsia. There is also a degree of uncertainty in the diagnostic criteria definitions. ^1,34^ Traditionally, most retrospective and modeling studies have used disease codes,^9^ which may be inaccurate.^35^ To improve the accuracy of the outcome, we developed and manually validated an algorithm based on structured data. In addition, as some patients utilized home blood pressure measurements, those values were not available in the EHR and, thus, were not included in the models. In the future, integrating home blood pressure monitoring may improve the predictive power of models. We also acknowledge that ACOG risk factors^1^ may not be an ideal comparative model to our models; however, this approach has been used by others,^10^ and most modeling studies have insufficient data to reproduce the results.^17^ Lastly, we used retrospective data to test and externally validate our models, and further prospective validation is needed to evaluate the true predictive power.

By using routinely collected data during scheduled prenatal visits, we demonstrate that accurate prediction of preeclampsia can be achieved. This design would allow risk assessment utilizing these models at every visit throughout pregnancy with updates in risk prediction, resulting in a more accurate ascertainment of individuals at risk who would benefit from prophylactic measures or increased surveillance. In the future, as better testing or prophylaxis become available, these AI tools can be used to select the group of individuals who would benefit the most.

## NON-STANDARD ABBREVIATIONS AND ACRONYMS

AUC: area under the receiver operating characteristic curve
BMI: body mass index
DBP: diastolic blood pressure
IUGR: intrauterine growth restriction
SBP: systolic blood pressure
SGA: small for gestational age
XGB: xgboost

## Contributions

B.E., V.K., and K.G. conceptualized and designed the study. B.E., R.C., and V.K. conducted all analyses, J.R. assisted with data processing. B.E. and V.K. drafted the manuscript, which was edited and critically reviewed by R.C., J.R., D.B., and K.G. Mass General Brigham was the source of data and location for all analyses. All authors had access to all study data. All authors read and approved the final manuscript and had final responsibility for the decision to submit it for publication.

## Data Sharing

The data used in this study are from Mass General Brigham patients and are not publicly available due to patient privacy and confidentiality.

## Acknowledgments.

KJG reports funding from NIH/NHLBI grants K08 HL146963, K08 HL146963-02S1, and R03 HL162756. VPK reports funding from the NIH/NHLBI grants 1K08HL161326-01A1, Anesthesia Patient Safety Foundation (APSF), and BWH IGNITE Award. The funders played no role in the study design, data collection, analysis, and interpretation of data, or the writing of this manuscript.

## REFERENCES

1. Gestational Hypertension and Preeclampsia: ACOG Practice Bulletin, Number 222. Obstet Gynecol 2020; 135(6): e237–e60.

2. Duley L. The global impact of pre-eclampsia and eclampsia. Semin Perinatol 2009; 33(3): 130–7.

3. Abalos E, Cuesta C, Grosso AL, Chou D, Say L. Global and regional estimates of preeclampsia and eclampsia: a systematic review. Eur J Obstet Gynecol Reprod Biol 2013; 170(1): 1–7.

4. Feng Y, Lian X, Guo K, Zhang G, Huang X. A comprehensive analysis of metabolomics and transcriptomics to reveal major metabolic pathways and potential biomarkers of human preeclampsia placenta. Front Genet 2022; 13: 1010657.

5. ACOG Committee Opinion No. 743: Low-Dose Aspirin Use During Pregnancy. Obstet Gynecol 2018; 132(1): e44–e52.

6. North RA, McCowan LM, Dekker GA, et al. Clinical risk prediction for pre-eclampsia in nulliparous women: development of model in international prospective cohort. BMJ 2011; 342: d1875.

7. Wright D, Syngelaki A, Akolekar R, Poon LC, Nicolaides KH. Competing risks model in screening for preeclampsia by maternal characteristics and medical history. Am J Obstet Gynecol 2015; 213(1): 62 e1–e10.

8. Wright D, Tan MY, O’Gorman N, et al. Predictive performance of the competing risk model in screening for preeclampsia. Am J Obstet Gynecol 2019; 220(2): 199 e1–e13.

9. Sandstrom A, Snowden JM, Bottai M, Stephansson O, Wikstrom AK. Routinely collected antenatal data for longitudinal prediction of preeclampsia in nulliparous women: a population-based study. Sci Rep 2021; 11(1): 17973.

10. Li S, Wang Z, Vieira LA, et al. Improving preeclampsia risk prediction by modeling pregnancy trajectories from routinely collected electronic medical record data. NPJ Digit Med 2022; 5(1): 68.

11. Maric I, Tsur A, Aghaeepour N, et al. Early prediction of preeclampsia via machine learning. Am J Obstet Gynecol MFM 2020; 2(2): 100100.

12. Kuklina EV, Whiteman MK, Hillis SD, et al. An enhanced method for identifying obstetric deliveries: implications for estimating maternal morbidity. Matern Child Health J 2008; 12(4): 469–77.

13. Cohen RY, Kovacheva VP. A Methodology for a Scalable, Collaborative, and Resource-Efficient Platform, MERLIN, to Facilitate Healthcare AI Research. IEEE J Biomed Health Inform 2023; 27(6): 3014–25.

14. American College of Obstetricians and Gynecologists’ Committee on Practice, Bulletins-Obstetrics. Gestational Hypertension and Preeclampsia: ACOG Practice Bulletin, Number 222. Obstet Gynecol 2020; 135(6): e237–e60.

15. NICE, National Institute for Health and Care Excellence. NICE guideline. Hypertension in pregnancy: diagnosis and management. 2019.

16. Bennett R, Mulla ZD, Parikh P, Hauspurg A, Razzaghi T. An imbalance-aware deep neural network for early prediction of preeclampsia. PLoS One 2022; 17(4): e0266042.

17. Snell KIE, Allotey J, Smuk M, et al. External validation of prognostic models predicting pre-eclampsia: individual participant data meta-analysis. BMC Med 2020; 18(1): 302.

18. Jhee JH, Lee S, Park Y, et al. Prediction model development of late-onset preeclampsia using machine learning-based methods. PLoS One 2019; 14(8): e0221202.

19. Ansbacher-Feldman Z, Syngelaki A, Meiri H, Cirkin R, Nicolaides KH, Louzoun Y. Machine-learning-based prediction of pre-eclampsia using first-trimester maternal characteristics and biomarkers. Ultrasound Obstet Gynecol 2022; 60(6): 739–45.

20. Akiba T SS, Yanase T, Ohta T, Koyama M. Optuna: A Next-generation Hyperparameter Optimization Framework. arXiv 2019; 1907.10902.

21. Obermeyer ZN, R. Stern, M. Eaneff, S. Bembeneck, E. Mullainathan, S. Algorithmic Bias Playbook: Center for Applied AI at Chicago Booth; 2021.

22. Lundberg S LS. A Unified Approach to Interpreting Model Predictions. arXiv 2017; 1705.07874.

23. Rolnik DL, Wright D, Poon LC, et al. Aspirin versus Placebo in Pregnancies at High Risk for Preterm Preeclampsia. N Engl J Med 2017; 377(7): 613–22.

24. Wu J, Li Y, Ma Y. Comparison of XGBoost and the Neural Network model on the class-balanced datasets. 2021 IEEE 3rd International Conference on Frontiers Technology of Information and Computer (ICFTIC); 2021 12-14 Nov. 2021; 2021. p. 457–61.

25. Hutcheon JA, Stephansson O, Cnattingius S, Bodnar LM, Wikstrom AK, Johansson K. Pregnancy Weight Gain Before Diagnosis and Risk of Preeclampsia: A Population-Based Cohort Study in Nulliparous Women. Hypertension 2018; 72(2): 433–41.

26. Elawad T, Scott G, Bone JN, et al. Risk factors for pre-eclampsia in clinical practice guidelines: Comparison with the evidence. BJOG: An International Journal of Obstetrics & Gynaecology; n/a(n/a).

27. Roell KR, Harmon QE, Klungsoyr K, Bauer AE, Magnus P, Engel SM. Clustering Longitudinal Blood Pressure Trajectories to Examine Heterogeneity in Outcomes Among Preeclampsia Cases and Controls. Hypertension 2021; 77(6): 2034–44.

28. Ananth CV, Keyes KM, Wapner RJ. Pre-eclampsia rates in the United States, 1980-2010: age-period-cohort analysis. BMJ 2013; 347: f6564.

29. Skjaerven R, Wilcox AJ, Lie RT. The interval between pregnancies and the risk of preeclampsia. N Engl J Med 2002; 346(1): 33–8.

30. Heilmann L, Siekmann U, Schmid-Schonbein H, Ludwig H. Hemoconcentration and pre-eclampsia. Arch Gynecol 1981; 231(1): 7–21.

31. Myatt L. The prediction of preeclampsia: the way forward. Am J Obstet Gynecol 2022; 226(2S): S1102–S7 e8.

32. Ghassemi M, Oakden-Rayner L, Beam AL. The false hope of current approaches to explainable artificial intelligence in health care. Lancet Digit Health 2021; 3(11): e745–e50.

33. Askie LM, Duley L, Henderson-Smart DJ, Stewart LA, Group PC. Antiplatelet agents for prevention of pre-eclampsia: a meta-analysis of individual patient data. Lancet 2007; 369(9575): 1791–8.

34. Reddy M, Fenn S, Rolnik DL, et al. The impact of the definition of preeclampsia on disease diagnosis and outcomes: a retrospective cohort study. Am J Obstet Gynecol 2021; 224(2): 217 e1–e11.

35. Labgold K, Stanhope KK, Joseph NT, Platner M, Jamieson DJ, Boulet SL. Validation of Hypertensive Disorders During Pregnancy: ICD-10 Codes in a High-burden Southeastern United States Hospital. Epidemiology 2021; 32(4): 591–7.

