## Supplemental Text, Figures and Tables for "An Interpretable Longitudinal Preeclampsia Risk Prediction Using Machine Learning"

### Supplementary Material – Table of Contents

Supplementary Methods........ 2
sTable 1. Hospitals providing data for the study........ 3
sTable 2. Summary characteristics of Cohort 1 ........ 4
sTable 3. Summary characteristics of Cohort 2 ........ 5
sTable 4. Comparison of Cohort 1 and Cohort 2........ 6
sTable 5. Summary characteristics of Cohort 1 at 14 weeks........ 8
sTable 6. Summary characteristics of Cohort 1 at 20 weeks........ 10
sTable 7. Summary characteristics of Cohort 1 at 24 weeks........12
sTable 8. Summary characteristics of Cohort 1 at 28 weeks........14
sTable 9. Summary characteristics of Cohort 1 at 32 weeks........16
sTable 10. Summary characteristics of Cohort 1 at 36 weeks........18
sTable 11. Summary characteristics of Cohort 1 at 39 weeks........20
sTable 12. Summary characteristics of Cohort 1 on admission........22
sTable 13. Cohort 1 Model Performance........24
Supplementary Results. External Validation ........25
sTable 14. Cohort 2 Model Performance........26
Results. Racial Bias Investigation ........27
sFigure 1. Performance in racial groups........28
Results. Individual patient predictions ........29
sFigure 2. Individual patient predictions........30
sFigure 3. Individual variables relationships........31
Supplementary Discussion........32
References ........33

### Supplementary Methods

#### External validation cohort

As data were available from eight hospitals, we developed all models using data from six hospitals (Cohort 1) and reserved the data from two hospitals (Cohort 2) for external validation. These two hospitals were chosen because they contain a representative sample of the entire population both in size and amount of data available.

#### Neural Networks

After processing the EHR to feature engineer, select relevant features, balance, and normalize the data, we fed the processed data into a fully connected deep neural network. We found that small changes to the network architecture could result in large changes in the results and thus used Optuna[1] and the built-in TPE Sampler to search the hyperparameter space and determine the best-performing model. Hyperparameters include the number of layers, number of nodes per layer, dropout rate, learning rate, batch size, and epochs. Skip connections were used to concatenate prior layers to ensure the passing of relevant information to subsequent layers.[2] RELU activation functions were applied between layers until the output layer, where we used a sigmoid function to produce a single risk score. The model was optimized using ADAM and a binary cross-entropy loss function.

Table S1. Data availability from all hospitals from the Mass General Brigham (MGB) Healthcare system.

| | **N (%)** |
|---|---|
| Brigham and Women's Hospital | 49606 (41.1) |
| Newton-Wellesley Hospital | 28021 (23.2) |
| Massachusetts General Hospital | 26134 (21.6) |
| Salem Hospital | 7550 (6.3) |
| Wentworth-Douglass Hospital | 4738 (3.9) |
| Cooley Dickinson Hospital | 2967 (2.5) |
| Martha Vineyard Hospital | 808 (0.7) |
| Nantucket Cottage Hospital | 764 (0.6) |
| Not Specified | 151 (0.1) |

Table S2. Summary characteristics of Cohort 1.

| | **Preeclampsia (n=6464)** | **No preeclampsia (n=106583)** | **P-value** |
|---|---|---|---|
| Maternal age at delivery, y | 33.04 (29.64 - 36.67) | 33.02 (30.01 - 35.92) | 0.12 |
| Self-reported race, White | 3825 (59.17%) | 71039 (66.65%) | < 0.01 |
| Self-reported race, Black | 1166 (18.04%) | 10283 (9.65%) | < 0.01 |
| Self-reported race, other | 1517 (23.47%) | 25798 (24.2%) | 0.24 |
| Self-reported ethnicity, Hispanic | 1263 (19.54%) | 16704 (15.67%) | < 0.01 |
| Self-reported ethnicity, non-Hispanic | 5201 (80.46%) | 89879 (84.33%) | < 0.01 |
| Tertiary hospital | 4712 (72.9%) | 71028 (66.64%) | < 0.01 |
| Community hospital | 1752 (27.1%) | 35555 (33.36%) | < 0.01 |
| Gravidity | 2.0 (1.0 - 3.0) | 2.0 (1.0 - 3.0) | 0.29 |
| Parity | 1.0 (0.0 - 1.75) | 1.0 (1.0 - 1.0) | < 0.01 |
| Gestational age at delivery, weeks | 37.57 (36.14 - 39.14) | 39.29 (38.57 - 40.14) | < 0.01 |
| BMI at delivery ($kg/m^2$) | 33.2 (29.2 - 38.4) | 29.9 (26.8 - 33.7) | < 0.01 |
| Maximal SBP during pregnancy, mmHg | 150.0 (140.0 - 160.0) | 129.0 (120.0 - 138.0) | < 0.01 |
| Maximal DBP during pregnancy, mmHg | 93.0 (86.0 - 99.0) | 80.0 (74.0 - 86.0) | < 0.01 |
| Family history of chronic hypertension | 2907 (44.97%) | 38883 (36.48%) | < 0.01 |
| Family history of preeclampsia | 144 (2.23%) | 2135 (2.0%) | 0.22 |
| Past history of chronic hypertension | 1868 (28.9%) | 5726 (5.37%) | < 0.01 |
| Past history of gestational hypertension | 500 (7.74%) | 3452 (3.24%) | < 0.01 |
| Past history of preeclampsia | 525 (8.12%) | 2179 (2.04%) | < 0.01 |
| Preterm delivery (<37 weeks) | 2143 (33.15%) | 7446 (6.99%) | < 0.01 |
| Antihypertensive medications | 4507 (69.72%) | 6044 (5.67%) | < 0.01 |
| Proteinuria | 3940 (60.95%) | 801 (0.75%) | < 0.01 |
| SGA or IUGR | 536 (8.29%) | 9241 (8.67%) | 0.32 |

Median (IQR) for continuous variables; n (%) for categorical variables; p-values for continuous variables based on Kruskal-Wallis rank sum test; for categorical variables based on Fisher's exact or Chi-squared test.
Abbreviations: SBP, systolic blood pressure; DBP, diastolic blood pressure, BMI, body mass index, SGA, small for gestational age; IUGR, intrauterine growth restriction.

Table S3. Summary characteristics of Cohort 2.

| | Preeclampsia (n=456) | No preeclampsia (n=7249) | P-value |
|---|---|---|---|
| Maternal age at delivery, y | 30.71 (27.18 - 34.4) | 31.19 (27.51 - 34.28) | 0.85 |
| Self-reported race, White | 415 (91.01%) | 6251 (86.23%) | 0.29 |
| Self-reported race, Black | 6 (1.32%) | 209 (2.88%) | 0.052 |
| Self-reported race, other | 36 (7.89%) | 828 (11.42%) | 0.029 |
| Self-reported ethnicity, Hispanic | 26 (5.7%) | 392 (5.41%) | 0.79 |
| Self-reported ethnicity, non-Hispanic | 430 (94.3%) | 6857 (94.59%) | 0.95 |
| Gravidity | 1.5 (1.0 - 3.0) | 2.0 (1.0 - 3.0) | 0.015 |
| Parity | 1.0 (0.0 - 1.0) | 1.0 (0.0 - 1.0) | < 0.01 |
| Gestational age at delivery, weeks | 38.57 (37.14 - 39.86) | 39.57 (39.0 - 40.43) | < 0.01 |
| BMI at delivery (kg/m$^2$) | 33.62 (29.43 - 39.02) | 31.2 (27.8 - 35.73) | < 0.01 |
| Maximal SBP during pregnancy, mmHg | 148.0 (141.0 - 158.0) | 130.0 (122.0 - 139.0) | < 0.01 |
| Maximal DBP during pregnancy, mmHg | 95.0 (88.0 - 101.0) | 82.0 (78.0 - 88.0) | < 0.01 |
| Family history of chronic hypertension | 168 (36.84%) | 2394 (33.03%) | 0.17 |
| Family history of preeclampsia | 5 (1.1%) | 27 (0.37%) | 0.02 |
| Past history of chronic hypertension | 28 (6.14%) | 128 (1.77%) | < 0.01 |
| Past history of gestational hypertension | 11 (2.41%) | 76 (1.05%) | < 0.01 |
| Past history of preeclampsia | 5 (1.1%) | 26 (0.36%) | 0.016 |
| Preterm delivery (<37 weeks), number of patients | 74 (16.23%) | 301 (4.15%) | < 0.01 |
| Antihypertensive medications | 260 (57.02%) | 311 (4.29%) | < 0.01 |
| Proteinuria | 346 (75.88%) | 85 (1.17%) | < 0.01 |
| SGA or IUGR | 43 (9.43%) | 522 (7.2%) | 0.088 |

Median (IQR) for continuous variables; n (%) for categorical variables; p-values for continuous variables based on Kruskal-Wallis rank sum test; for categorical variables based on Fisher's exact or Chi-squared test.
Abbreviations: SBP, systolic blood pressure; DBP, diastolic blood pressure, BMI, body mass index, SGA, small for gestational age; IUGR, intrauterine growth restriction.

Table S4. Comparison of Cohort 1 and Cohort 2.

| | **Cohort 1 (n=113,047)** | **Cohort 2 (n=7,705)** | **P-Value** |
|---|---|---|---|
| Maternal age at delivery, y | 33.02 (29.99 - 35.96) | 31.16 (27.49 - 34.3) | < 0.01 |
| Self-reported race, White | 74864 (66.22%) | 6666 (86.52%) | < 0.01 |
| Self-reported race, Black | 11449 (10.13%) | 215 (2.79%) | < 0.01 |
| Self-reported race, other | 27315 (24.16%) | 864 (11.21%) | < 0.01 |
| Self-reported ethnicity, Hispanic | 17967 (15.89%) | 418 (5.43%) | < 0.01 |
| Self-reported ethnicity, non-Hispanic | 95080 (84.11%) | 7287 (94.57%) | < 0.01 |
| Tertiary hospital | 75740 (67.0%) | 0 (0.0%) | < 0.01 |
| Community hospital | 37307 (33.0%) | 7705 (100.0%) | < 0.01 |
| Alcohol use | 29334 (25.95%) | 1167 (15.15%) | < 0.01 |
| Smoking | 11542 (10.21%) | 939 (12.19%) | < 0.01 |
| Illicit drugs | 4925 (4.36%) | 727 (9.44%) | < 0.01 |
| Gravidity | 2.0 (1.0 - 3.0) | 2.0 (1.0 - 3.0) | < 0.01 |
| Parity | 1.0 (1.0 - 1.0) | 1.0 (0.0 - 1.0) | 0.063 |
| Nulliparous | 14973 (13.24%) | 1016 (13.19%) | 0.89 |
| Gestational age at delivery, weeks | 39.29 (38.43 - 40.14) | 39.43 (38.86 - 40.43) | < 0.01 |
| Interpregnancy interval, y | 2.32 (1.83 - 3.04) | 2.0 (1.62 - 2.5) | < 0.01 |
| Maximal SBP during pregnancy, mmHg | 130.0 (121.0 - 140.0) | 130.0 (122.0 - 141.0) | 0.015 |
| Maximal DBP during pregnancy, mmHg | 80.0 (75.0 - 87.0) | 82.0 (78.0 - 90.0) | < 0.01 |
| Maximal heart rate during pregnancy, bpm | 95.0 (85.0 - 105.0) | 96.0 (86.0 - 107.0) | < 0.01 |
| Body mass index at delivery (kg/m$^2$) | 30.0 (26.9 - 34.0) | 31.3 (27.9 - 36.0) | < 0.01 |
| Weight at delivery, kg | 79.83 (71.49 - 90.72) | 83.91 (73.94 - 96.89) | < 0.01 |
| Family history of chronic hypertension | 41790 (36.97%) | 2562 (33.25%) | < 0.01 |
| Family history of heart disease | 1891 (1.67%) | 46 (0.6%) | < 0.01 |
| Family history of preeclampsia | 2279 (2.02%) | 32 (0.42%) | < 0.01 |
| Past history of chronic hypertension | 7594 (6.72%) | 156 (2.02%) | < 0.01 |
| Past history of gestational hypertension | 3952 (3.5%) | 87 (1.13%) | < 0.01 |
| Past history of preeclampsia | 2704 (2.39%) | 31 (0.4%) | < 0.01 |
| Past history of diabetes | 4978 (4.4%) | 261 (3.39%) | < 0.01 |
| Gestational hypertension | 12328 (10.91%) | 600 (7.79%) | < 0.01 |
| Anemia during pregnancy | 18084 (16.0%) | 395 (5.13%) | < 0.01 |
| Headaches during pregnancy | 13973 (12.36%) | 277 (3.6%) | < 0.01 |
| Autoimmune history | 5537 (4.9%) | 163 (2.12%) | < 0.01 |
| High risk pregnancy | 57251 (50.64%) | 2822 (36.63%) | < 0.01 |
| Gestational diabetes | 14649 (12.96%) | 486 (6.31%) | < 0.01 |
| Baby oligohydramnios | 2346 (2.08%) | 58 (0.75%) | < 0.01 |
| Preterm delivery (<37 weeks) | 9589 (8.48%) | 375 (4.87%) | < 0.01 |
| Hyperemesis gravidarum | 8992 (7.95%) | 152 (1.97%) | < 0.01 |
| Pregnancy related fatigue | 21030 (18.6%) | 1604 (20.82%) | < 0.01 |
| Preeclampsia | 6464 (5.72%) | 456 (5.92%) | 0.48 |
| Maximal lactate dehydrogenase during pregnancy | 206.0 (176.0 - 251.0) | 188.0 (163.0 - 228.0) | < 0.01 |
| Maximal PT/INR during pregnancy | 1.0 (1.0 - 1.1) | 1.0 (1.0 - 1.1) | 0.13 |
| Proteinuria | 4741 (4.19%) | 431 (5.59%) | < 0.01 |
| Maximal AST during pregnancy | 20.0 (17.0 - 27.0) | 19.0 (15.0 - 26.0) | 0.024 |
| Maximal uric acid during pregnancy | 4.8 (4.0 - 5.8) | 4.8 (4.0 - 5.8) | 0.71 |
| Maximal white blood count during pregnancy | 9.44 (8.67 - 9.94) | 9.59 (8.76 - 12.44) | < 0.01 |
| Maximal ALT during pregnancy | 15.0 (10.0 - 24.0) | 18.0 (13.0 - 27.0) | < 0.01 |
| Maximal serum calcium during pregnancy | 9.3 (9.1 - 9.6) | 9.1 (8.8 - 9.4) | < 0.01 |
| Maximal serum creatinine during pregnancy | 0.64 (0.56 - 0.74) | 0.6 (0.51 - 0.7) | < 0.01 |
| Maximal eosinophils during pregnancy | 1.2 (0.7 - 2.0) | 1.1 (0.6 - 1.8) | 0.078 |
| Maximal serum glucose during pregnancy | 89.0 (81.0 - 96.0) | 90.0 (83.0 - 97.0) | < 0.01 |
| Minimal hemoglobin during pregnancy | 11.1 (10.5 - 11.7) | 11.1 (10.5 - 11.8) | < 0.01 |
| Maximal lymphocytes during pregnancy | 22.6 (18.4 - 27.2) | 18.0 (15.0 - 23.0) | < 0.01 |
| Maximal platelets during pregnancy | 257.0 (218.0 - 301.0) | 254.0 (215.0 - 299.0) | 0.22 |
| Minimal red blood cell count during pregnancy | 3.59 (3.28 - 3.86) | 3.57 (3.23 - 3.86) | < 0.01 |
| Antihypertensive medications | 10551 (9.33%) | 571 (7.41%) | < 0.01 |
| SGA or IUGR | 9777 (8.65%) | 565 (7.33%) | < 0.01 |

Median (IQR) for continuous variables; n (%) for categorical variables; p-values for continuous variables based on Kruskal-Wallis rank sum test; for categorical variables based on Fisher's exact or Chi-squared test.

Abbreviations: SBP, systolic blood pressure; DBP, diastolic blood pressure, BMI, body mass index, PT/INR, prothrombin time/international normalized ratio; AST, aspartate transferase; ALT, alanine transaminase; SGA, small for gestational age; IUGR, intrauterine growth restriction.

Table S5. Summary characteristics of Cohort 1 patient data at 14 weeks gestation.

| | **Preeclampsia (n=6464)** | **No preeclampsia (n=106581)** | **P-Value** |
|---|---|---|---|
| Maternal age, y | 33.03 (29.66 - 36.65) | 32.99 (30.03 - 35.9) | 0.12 |
| Self-reported race, White | 3825 (59.17%) | 71037 (66.65%) | < 0.01 |
| Self-reported race, Black | 1166 (18.04%) | 10283 (9.65%) | < 0.01 |
| Self-reported race, other | 1517 (23.47%) | 25798 (24.2%) | 0.24 |
| Self-reported ethnicity, Hispanic | 1263 (19.54%) | 16704 (15.67%) | < 0.01 |
| Self-reported ethnicity, non-Hispanic | 5201 (80.46%) | 89879 (84.33%) | < 0.01 |
| Private insurance | 5085 (78.67%) | 90488 (84.9%) | < 0.01 |
| Public insurance | 1851 (28.64%) | 22076 (20.71%) | < 0.01 |
| Alcohol use history | 1577 (24.4%) | 24164 (22.67%) | < 0.01 |
| Smoking history | 877 (13.57%) | 10403 (9.76%) | < 0.01 |
| Illicit drugs history | 317 (4.9%) | 3213 (3.01%) | < 0.01 |
| Gravidity | 2.0 (1.0 - 3.0) | 2.0 (1.0 - 3.0) | 0.61 |
| Parity | 1.0 (1.0 - 1.0) | 1.0 (1.0 - 1.0) | < 0.01 |
| In vitro fertilization | 396 (6.13%) | 4141 (3.89%) | < 0.01 |
| Nulliparous | 1183 (18.3%) | 13790 (12.94%) | < 0.01 |
| Interpregnancy interval, y | 2.3 (2.3 - 2.3) | 2.3 (2.3 - 2.3) | 0.93 |
| Multiple gestation | 479 (7.41%) | 2870 (2.69%) | < 0.01 |
| Maximal SBP 0-14 weeks, mmHg | 120.0 (118.0 - 130.0) | 118.0 (112.0 - 120.0) | < 0.01 |
| Maximal DBP 0-14 weeks, mmHg | 72.0 (71.0 - 81.0) | 71.0 (68.0 - 74.0) | < 0.01 |
| Maximal heart rate 0-14 weeks, bpm | 85.0 (85.0 - 85.0) | 85.0 (85.0 - 85.0) | < 0.01 |
| Maximal body mass index 0-14 weeks ($kg/m^2$) | 25.1 (25.1 - 30.5) | 25.1 (23.8 - 26.0) | < 0.01 |
| Maximal weight 0-14 weeks, kg | 67.31 (67.31 - 82.49) | 67.31 (63.05 - 70.85) | < 0.01 |
| Family history of chronic hypertension | 2907 (44.97%) | 38881 (36.48%) | < 0.01 |
| Family history of heart disease | 2980 (46.1%) | 40548 (38.04%) | < 0.01 |
| Family history of preeclampsia | 144 (2.23%) | 2135 (2.0%) | 0.22 |
| Family history of diabetes | 329 (5.09%) | 4078 (3.83%) | < 0.01 |
| Family history of hyperlipidemia | 120 (1.86%) | 1571 (1.47%) | 0.015 |
| Family history of stroke | 157 (2.43%) | 1854 (1.74%) | < 0.01 |
| Past history of diabetes | 507 (7.84%) | 3036 (2.85%) | < 0.01 |
| Past history of gestational diabetes | 359 (5.55%) | 4260 (4.0%) | < 0.01 |
| Past history of cesarean delivery | 1458 (22.56%) | 19675 (18.46%) | < 0.01 |
| Past history of preterm birth | 510 (7.89%) | 4582 (4.3%) | < 0.01 |
| Past history of gynecologic surgery | 780 (12.07%) | 13528 (12.69%) | 0.17 |
| Past history of asthma | 949 (14.68%) | 11136 (10.45%) | < 0.01 |
| Past history of chronic hypertension | 1868 (28.9%) | 5725 (5.37%) | < 0.01 |
| Past history of gestational hypertension | 500 (7.74%) | 3451 (3.24%) | < 0.01 |
| Past history of high-risk pregnancy | 1166 (18.04%) | 18472 (17.33%) | 0.19 |
| Past history of hyperemesis gravidarum | 1037 (16.04%) | 17344 (16.27%) | 0.66 |
| Past history of migraine | 900 (13.92%) | 10867 (10.2%) | < 0.01 |
| Past history of obesity | 2050 (31.71%) | 16482 (15.46%) | < 0.01 |
| Past history of preeclampsia | 525 (8.12%) | 2179 (2.04%) | < 0.01 |
| Past history of pregnancy related fatigue | 522 (8.08%) | 9311 (8.74%) | 0.08 |
| Past history of sexually transmitted disease | 345 (5.34%) | 5163 (4.84%) | 0.081 |
| Chronic hypertension | 810 (12.53%) | 2312 (2.17%) | < 0.01 |
| Anemia during pregnancy | 715 (11.06%) | 10315 (9.68%) | < 0.01 |
| Headaches during pregnancy | 950 (14.7%) | 10861 (10.19%) | < 0.01 |
| Autoimmune disease | 635 (9.82%) | 4902 (4.6%) | < 0.01 |
| High risk pregnancy | 2806 (43.41%) | 37505 (35.19%) | < 0.01 |
| Hyperemesis gravidarum | 304 (4.7%) | 3848 (3.61%) | < 0.01 |
| Pregnancy related fatigue | 474 (7.33%) | 6917 (6.49%) | 0.01 |
| Proteinuria 0-14 weeks | 69 (1.07%) | 37 (0.03%) | < 0.01 |
| Maximal AST 0-14 weeks | 17.0 (17.0 - 17.0) | 17.0 (17.0 - 17.0) | < 0.01 |
| Maximal white blood count 0-14 weeks | 8.51 (8.1 - 9.62) | 8.51 (7.9 - 9.2) | < 0.01 |
| Maximal ALT 0-14 weeks | 15.0 (15.0 - 15.0) | 15.0 (15.0 - 15.0) | < 0.01 |
| Maximal serum calcium 0-14 weeks | 9.3 (9.3 - 9.3) | 9.3 (9.3 - 9.3) | < 0.01 |
| Maximal serum creatinine 0-14 weeks | 0.62 (0.62 - 0.62) | 0.62 (0.62 - 0.62) | < 0.01 |
| Maximal eosinophils 0-14 weeks | 1.0 (1.0 - 1.0) | 1.0 (1.0 - 1.0) | 0.089 |
| Maximal serum glucose 0-14 weeks | 89.0 (89.0 - 89.0) | 89.0 (89.0 - 89.0) | < 0.01 |
| Minimal hemoglobin 0-14 weeks | 12.7 (12.2 - 13.0) | 12.7 (12.3 - 13.0) | 0.082 |
| Maximal lymphocytes 0-14 weeks | 23.4 (23.4 - 23.4) | 23.4 (23.4 - 23.4) | 0.28 |
| Maximal platelets 0-14 weeks | 258.0 (250.0 - 298.0) | 258.0 (239.0 - 276.0) | < 0.01 |
| Minimal red blood cell count 0-14 weeks | 4.24 (4.14 - 4.42) | 4.24 (4.12 - 4.34) | < 0.01 |
| Antihypertensive medications 0-14 weeks | 40 (0.62%) | 26 (0.02%) | < 0.01 |

Median (IQR) for continuous variables; n (%) for categorical variables; p-values for continuous variables based on Kruskal-Wallis rank sum test; for categorical variables based on Fisher's exact or Chi-squared test.

Abbreviations: SBP, systolic blood pressure; DBP, diastolic blood pressure, BMI, body mass index, PT/INR, prothrombin time/international normalized ratio; AST, aspartate transferase; ALT, alanine transaminase; SGA, small for gestational age; IUGR, intrauterine growth restriction.

Table S6. Summary characteristics of Cohort 1 patient data at 20 weeks gestation.

| | **Preeclampsia (n=6464)** | **No preeclampsia (n=106581)** | **P-Value** |
|---|---|---|---|
| Maternal age, y | 33.03 (29.66 - 36.65) | 32.99 (30.03 - 35.9) | 0.12 |
| Self-reported race, White | 3825 (59.17%) | 71037 (66.65%) | < 0.01 |
| Self-reported race, Black | 1166 (18.04%) | 10283 (9.65%) | < 0.01 |
| Self-reported race, other | 1517 (23.47%) | 25798 (24.2%) | 0.24 |
| Self-reported ethnicity, Hispanic | 1263 (19.54%) | 16704 (15.67%) | < 0.01 |
| Self-reported ethnicity, non-Hispanic | 5201 (80.46%) | 89879 (84.33%) | < 0.01 |
| Private insurance | 5085 (78.67%) | 90488 (84.9%) | < 0.01 |
| Public insurance | 1851 (28.64%) | 22076 (20.71%) | < 0.01 |
| Alcohol use history | 1577 (24.4%) | 24164 (22.67%) | < 0.01 |
| Smoking history | 877 (13.57%) | 10403 (9.76%) | < 0.01 |
| Illicit drugs history | 317 (4.9%) | 3213 (3.01%) | < 0.01 |
| Gravidity | 2.0 (1.0 - 3.0) | 2.0 (1.0 - 3.0) | 0.61 |
| Parity | 1.0 (1.0 - 1.0) | 1.0 (1.0 - 1.0) | < 0.01 |
| In vitro fertilization | 396 (6.13%) | 4141 (3.89%) | < 0.01 |
| Nulliparous | 1183 (18.3%) | 13790 (12.94%) | < 0.01 |
| Interpregnancy interval, y | 2.3 (2.3 - 2.3) | 2.3 (2.3 - 2.3) | 0.93 |
| Multiple gestation | 479 (7.41%) | 2870 (2.69%) | < 0.01 |
| Maximal SBP 0-20 weeks, mmHg | 122.0 (120.0 - 135.0) | 120.0 (113.0 - 122.0) | < 0.01 |
| Maximal DBP 0-20 weeks, mmHg | 76.0 (72.0 - 83.0) | 72.0 (70.0 - 77.0) | < 0.01 |
| Maximal heart rate 0-20 weeks, bpm | 87.0 (87.0 - 87.0) | 87.0 (87.0 - 87.0) | < 0.01 |
| Maximal body mass index 0-20 weeks (kg/m$^2$) | 25.9 (25.9 - 31.9) | 25.9 (24.3 - 27.2) | < 0.01 |
| Maximal weight 0-20 weeks, kg | 69.4 (69.12 - 86.18) | 69.4 (64.41 - 74.0) | < 0.01 |
| Family history of chronic hypertension | 2907 (44.97%) | 38881 (36.48%) | < 0.01 |
| Family history of heart disease | 2980 (46.1%) | 40548 (38.04%) | < 0.01 |
| Family history of preeclampsia | 144 (2.23%) | 2135 (2.0%) | 0.22 |
| Family history of diabetes | 329 (5.09%) | 4078 (3.83%) | < 0.01 |
| Family history of hyperlipidemia | 120 (1.86%) | 1571 (1.47%) | 0.015 |
| Family history of stroke | 157 (2.43%) | 1854 (1.74%) | < 0.01 |
| Past history of diabetes | 507 (7.84%) | 3036 (2.85%) | < 0.01 |
| Past history of cesarean delivery | 1458 (22.56%) | 19675 (18.46%) | < 0.01 |
| Past history of preterm birth | 510 (7.89%) | 4582 (4.3%) | < 0.01 |
| Past history of gynecologic surgery | 780 (12.07%) | 13528 (12.69%) | 0.17 |
| Past history of asthma | 949 (14.68%) | 11136 (10.45%) | < 0.01 |
| Past history of chronic hypertension | 1868 (28.9%) | 5725 (5.37%) | < 0.01 |
| Past history of gestational hypertension | 500 (7.74%) | 3451 (3.24%) | < 0.01 |
| Past history of high-risk pregnancy | 1166 (18.04%) | 18472 (17.33%) | 0.19 |
| Past history of hyperemesis gravidarum | 1037 (16.04%) | 17344 (16.27%) | 0.66 |
| Past history of migraine | 900 (13.92%) | 10867 (10.2%) | < 0.01 |
| Past history of obesity | 2050 (31.71%) | 16482 (15.46%) | < 0.01 |
| Past history of preeclampsia | 525 (8.12%) | 2179 (2.04%) | < 0.01 |
| Past history of pregnancy related fatigue | 522 (8.08%) | 9311 (8.74%) | 0.08 |
| Past history of sexually transmitted disease | 345 (5.34%) | 5163 (4.84%) | 0.081 |
| Chronic hypertension | 942 (14.57%) | 2673 (2.51%) | < 0.01 |
| Gestational diabetes | 420 (6.5%) | 4612 (4.33%) | < 0.01 |
| Anemia during pregnancy | 767 (11.87%) | 10805 (10.14%) | < 0.01 |
| Headaches during pregnancy | 976 (15.1%) | 11245 (10.55%) | < 0.01 |
| Autoimmune disease | 635 (9.82%) | 4902 (4.6%) | < 0.01 |
| High risk pregnancy | 3099 (47.94%) | 41091 (38.55%) | < 0.01 |
| Hyperemesis gravidarum | 402 (6.22%) | 5166 (4.85%) | < 0.01 |
| Pregnancy related fatigue | 682 (10.55%) | 9591 (9.0%) | 0.01 |
| Proteinuria 0-20 weeks | 126 (1.95%) | 69 (0.06%) | < 0.01 |
| Maximal AST 0-20 weeks | 17.0 (17.0 - 17.0) | 17.0 (17.0 - 17.0) | < 0.01 |
| Maximal white blood count 0-20 weeks | 8.57 (8.1 - 9.73) | 8.57 (7.88 - 9.33) | < 0.01 |
| Maximal ALT 0-20 weeks | 15.0 (15.0 - 15.0) | 15.0 (15.0 - 15.0) | < 0.01 |
| Maximal serum calcium 0-20 weeks | 9.3 (9.3 - 9.3) | 9.3 (9.3 - 9.3) | < 0.01 |
| Maximal serum creatinine 0-20 weeks | 0.6 (0.6 - 0.6) | 0.6 (0.6 - 0.6) | < 0.01 |
| Maximal eosinophils 0-20 weeks | 1.0 (1.0 - 1.0) | 1.0 (1.0 - 1.0) | 0.037 |
| Maximal serum glucose 0-20 weeks | 89.0 (89.0 - 89.0) | 89.0 (89.0 - 89.0) | < 0.01 |
| Minimal hemoglobin 0-20 weeks | 12.6 (12.0 - 12.9) | 12.6 (12.2 - 12.9) | 0.028 |
| Maximal lymphocytes 0-20 weeks | 23.2 (23.2 - 23.2) | 23.2 (23.2 - 23.2) | 0.58 |
| Maximal platelets 0-20 weeks | 258.0 (247.0 - 300.0) | 258.0 (236.0 - 278.0) | < 0.01 |
| Minimal red blood cell count 0-20 weeks | 4.21 (4.08 - 4.41) | 4.21 (4.08 - 4.33) | < 0.01 |
| Antihypertensive medications 0-20 weeks | 74 (1.14%) | 51 (0.05%) | < 0.01 |

Median (IQR) for continuous variables; n (%) for categorical variables; p-values for continuous variables based on Kruskal-Wallis rank sum test; for categorical variables based on Fisher's exact or Chi-squared test.
Abbreviations: SBP, systolic blood pressure; DBP, diastolic blood pressure, BMI, body mass index, PT/INR, prothrombin time/international normalized ratio; AST, aspartate transferase; ALT, alanine transaminase; SGA, small for gestational age; IUGR, intrauterine growth restriction.

Table S7. Summary characteristics of Cohort 1 patient data at 24 weeks gestation.

| | **Preeclampsia (n=6464)** | **No preeclampsia (n=106581)** | **P-Value** |
|---|---|---|---|
| Maternal age, y | 33.03 (29.66 - 36.65) | 32.99 (30.03 - 35.9) | 0.12 |
| Self-reported race, White | 3825 (59.17%) | 71037 (66.65%) | < 0.01 |
| Self-reported race, Black | 1166 (18.04%) | 10283 (9.65%) | < 0.01 |
| Self-reported race, other | 1517 (23.47%) | 25798 (24.2%) | 0.24 |
| Self-reported ethnicity, Hispanic | 1263 (19.54%) | 16704 (15.67%) | < 0.01 |
| Self-reported ethnicity, non-Hispanic | 5201 (80.46%) | 89879 (84.33%) | < 0.01 |
| Private insurance | 5085 (78.67%) | 90488 (84.9%) | < 0.01 |
| Public insurance | 1851 (28.64%) | 22076 (20.71%) | < 0.01 |
| Alcohol use history | 1577 (24.4%) | 24164 (22.67%) | < 0.01 |
| Smoking history | 877 (13.57%) | 10403 (9.76%) | < 0.01 |
| Illicit drugs history | 317 (4.9%) | 3213 (3.01%) | < 0.01 |
| Gravidity | 2.0 (1.0 - 3.0) | 2.0 (1.0 - 3.0) | 0.61 |
| Parity | 1.0 (1.0 - 1.0) | 1.0 (1.0 - 1.0) | < 0.01 |
| In vitro fertilization | 396 (6.13%) | 4141 (3.89%) | < 0.01 |
| Nulliparous | 1183 (18.3%) | 13790 (12.94%) | < 0.01 |
| Interpregnancy interval, y | 2.3 (2.3 - 2.3) | 2.3 (2.3 - 2.3) | 0.93 |
| Multiple gestation | 479 (7.41%) | 2870 (2.69%) | < 0.01 |
| Maximal SBP 0-24 weeks, mmHg | 124.0 (120.0 - 136.0) | 120.0 (114.0 - 123.0) | < 0.01 |
| Maximal DBP 0-24 weeks, mmHg | 78.0 (73.0 - 84.0) | 73.0 (70.0 - 78.0) | < 0.01 |
| Maximal heart rate 0-24 weeks, bpm | 88.0 (88.0 - 89.0) | 88.0 (88.0 - 88.0) | < 0.01 |
| Maximal body mass index 0-24 weeks, ($kg/m^2$) | 26.62 (26.6 - 32.8) | 26.6 (24.9 - 28.1) | < 0.01 |
| Maximal weight 0-24 weeks, kg | 71.67 (70.74 - 88.38) | 71.21 (65.95 - 76.29) | < 0.01 |
| Family history of chronic hypertension | 2907 (44.97%) | 38881 (36.48%) | < 0.01 |
| Family history of heart disease | 2980 (46.1%) | 40548 (38.04%) | < 0.01 |
| Family history of preeclampsia | 144 (2.23%) | 2135 (2.0%) | 0.22 |
| Family history of diabetes | 329 (5.09%) | 4078 (3.83%) | < 0.01 |
| Family history of hyperlipidemia | 120 (1.86%) | 1571 (1.47%) | 0.015 |
| Family history of stroke | 157 (2.43%) | 1854 (1.74%) | < 0.01 |
| Past history of diabetes | 507 (7.84%) | 3036 (2.85%) | < 0.01 |
| Past history of cesarean delivery | 1458 (22.56%) | 19675 (18.46%) | < 0.01 |
| Past history of preterm birth | 510 (7.89%) | 4582 (4.3%) | < 0.01 |
| Past history of gynecologic surgery | 780 (12.07%) | 13528 (12.69%) | 0.17 |
| Past history of asthma | 949 (14.68%) | 11136 (10.45%) | < 0.01 |
| Past history of chronic hypertension | 1868 (28.9%) | 5725 (5.37%) | < 0.01 |
| Past history of gestational hypertension | 500 (7.74%) | 3451 (3.24%) | < 0.01 |
| Past history of high-risk pregnancy | 1166 (18.04%) | 18472 (17.33%) | 0.19 |
| Past history of hyperemesis gravidarum | 1037 (16.04%) | 17344 (16.27%) | 0.66 |
| Past history of migraine | 900 (13.92%) | 10867 (10.2%) | < 0.01 |
| Past history of obesity | 2050 (31.71%) | 16482 (15.46%) | < 0.01 |
| Past history of preeclampsia | 525 (8.12%) | 2179 (2.04%) | < 0.01 |
| Past history of pregnancy related fatigue | 522 (8.08%) | 9311 (8.74%) | 0.08 |
| Past history of sexually transmitted disease | 345 (5.34%) | 5163 (4.84%) | 0.081 |
| Chronic or gestational hypertension | 1045 (16.17%) | 2943 (2.76%) | < 0.01 |
| Gestational diabetes | 449 (6.95%) | 4936 (4.63%) | < 0.01 |
| Anemia during pregnancy | 789 (12.21%) | 11097 (10.41%) | < 0.01 |
| Headaches during pregnancy | 992 (15.35%) | 11383 (10.68%) | < 0.01 |
| Autoimmune disease | 635 (9.82%) | 4902 (4.6%) | < 0.01 |
| High risk pregnancy | 3255 (50.36%) | 43013 (40.36%) | < 0.01 |
| Hyperemesis gravidarum | 445 (6.88%) | 5960 (5.59%) | < 0.01 |
| Pregnancy related fatigue | 793 (12.27%) | 11426 (10.72%) | < 0.01 |
| Proteinuria 0-24 weeks, | 151 (2.34%) | 78 (0.07%) | < 0.01 |
| Maximal AST 0-24 weeks, | 17.0 (17.0 - 17.0) | 17.0 (17.0 - 17.0) | < 0.01 |
| Maximal white blood count 0-24 weeks | 8.63 (8.14 - 9.8) | 8.63 (7.9 - 9.39) | < 0.01 |
| Maximal ALT 0-24 weeks | 15.0 (15.0 - 15.0) | 15.0 (15.0 - 15.0) | < 0.01 |
| Maximal serum calcium 0-24 weeks | 9.3 (9.3 - 9.3) | 9.3 (9.3 - 9.3) | < 0.01 |
| Maximal serum creatinine 0-24 weeks | 0.6 (0.6 - 0.6) | 0.6 (0.6 - 0.6) | < 0.01 |
| Maximal eosinophils 0-24 weeks | 1.1 (1.1 - 1.1) | 1.1 (1.1 - 1.1) | 0.036 |
| Maximal serum glucose 0-24 weeks | 89.0 (89.0 - 89.0) | 89.0 (89.0 - 89.0) | < 0.01 |
| Minimal hemoglobin 0-24 weeks | 12.5 (11.9 - 12.9) | 12.5 (12.1 - 12.9) | < 0.01 |
| Maximal lymphocytes 0-24 weeks | 23.1 (23.1 - 23.1) | 23.1 (23.1 - 23.1) | 0.84 |
| Maximal platelets 0-24 weeks | 258.0 (246.0 - 302.0) | 258.0 (236.0 - 279.0) | < 0.01 |
| Minimal red blood cell count 0-24 weeks | 4.18 (4.03 - 4.38) | 4.18 (4.05 - 4.32) | 0.19 |
| Antihypertensive medications 0-24 weeks | 107 (1.66%) | 71 (0.07%) | < 0.01 |

Median (IQR) for continuous variables; n (%) for categorical variables; p-values for continuous variables based on Kruskal-Wallis rank sum test; for categorical variables based on Fisher's exact or Chi-squared test.
Abbreviations: SBP, systolic blood pressure; DBP, diastolic blood pressure, BMI, body mass index, PT/INR, prothrombin time/international normalized ratio; AST, aspartate transferase; ALT, alanine transaminase; SGA, small for gestational age; IUGR, intrauterine growth restriction.

Table S8. Summary characteristics of Cohort 1 patient data at 28 weeks gestation.

| | **Preeclampsia (n=6464)** | **No preeclampsia (n=106581)** | **P-Value** |
|---|---|---|---|
| Maternal age, y | 33.03 (29.66 - 36.65) | 32.99 (30.03 - 35.9) | 0.12 |
| Self-reported race, White | 3825 (59.17%) | 71037 (66.65%) | < 0.01 |
| Self-reported race, Black | 1166 (18.04%) | 10283 (9.65%) | < 0.01 |
| Self-reported race, other | 1517 (23.47%) | 25798 (24.2%) | 0.24 |
| Self-reported ethnicity, Hispanic | 1263 (19.54%) | 16704 (15.67%) | < 0.01 |
| Self-reported ethnicity, non-Hispanic | 5201 (80.46%) | 89879 (84.33%) | < 0.01 |
| Private insurance | 5085 (78.67%) | 90488 (84.9%) | < 0.01 |
| Public insurance | 1851 (28.64%) | 22076 (20.71%) | < 0.01 |
| Alcohol use history | 1577 (24.4%) | 24164 (22.67%) | < 0.01 |
| Smoking history | 877 (13.57%) | 10403 (9.76%) | < 0.01 |
| Illicit drugs history | 317 (4.9%) | 3213 (3.01%) | < 0.01 |
| Gravidity | 2.0 (1.0 - 3.0) | 2.0 (1.0 - 3.0) | 0.61 |
| Parity | 1.0 (1.0 - 1.0) | 1.0 (1.0 - 1.0) | < 0.01 |
| In vitro fertilization | 396 (6.13%) | 4141 (3.89%) | < 0.01 |
| Nulliparous | 1183 (18.3%) | 13790 (12.94%) | < 0.01 |
| Interpregnancy interval, y | 2.3 (2.3 - 2.3) | 2.3 (2.3 - 2.3) | 0.93 |
| Multiple gestation | 479 (7.41%) | 2870 (2.69%) | < 0.01 |
| Maximal SBP 0-28 weeks, mmHg | 126.0 (120.0 - 139.0) | 120.0 (116.0 - 124.0) | < 0.01 |
| Maximal DBP 0-28 weeks, mmHg | 80.0 (74.0 - 86.0) | 74.0 (70.0 - 78.0) | < 0.01 |
| Maximal heart rate 0-28 weeks, bpm | 89.0 (89.0 - 92.0) | 89.0 (89.0 - 89.0) | < 0.01 |
| Maximal body mass index 0-28 weeks, (kg/m$^2$) | 28.0 (27.5 - 33.8) | 27.5 (25.7 - 29.2) | < 0.01 |
| Maximal weight 0-28 weeks, kg | 75.18 (72.57 - 91.17) | 73.71 (68.04 - 79.29) | < 0.01 |
| Family history of chronic hypertension | 2907 (44.97%) | 38881 (36.48%) | < 0.01 |
| Family history of heart disease | 2980 (46.1%) | 40548 (38.04%) | < 0.01 |
| Family history of preeclampsia | 144 (2.23%) | 2135 (2.0%) | 0.22 |
| Family history of diabetes | 329 (5.09%) | 4078 (3.83%) | < 0.01 |
| Family history of hyperlipidemia | 120 (1.86%) | 1571 (1.47%) | 0.015 |
| Family history of stroke | 157 (2.43%) | 1854 (1.74%) | < 0.01 |
| Past history of diabetes | 507 (7.84%) | 3036 (2.85%) | < 0.01 |
| Past history of cesarean delivery | 1458 (22.56%) | 19675 (18.46%) | < 0.01 |
| Past history of preterm birth | 510 (7.89%) | 4582 (4.3%) | < 0.01 |
| Past history of gynecologic surgery | 780 (12.07%) | 13528 (12.69%) | 0.17 |
| Past history of asthma | 949 (14.68%) | 11136 (10.45%) | < 0.01 |
| Past history of chronic hypertension | 1868 (28.9%) | 5725 (5.37%) | < 0.01 |
| Past history of gestational hypertension | 500 (7.74%) | 3451 (3.24%) | < 0.01 |
| Past history of high-risk pregnancy | 1166 (18.04%) | 18472 (17.33%) | 0.19 |
| Past history of hyperemesis gravidarum | 1037 (16.04%) | 17344 (16.27%) | 0.66 |
| Past history of migraine | 900 (13.92%) | 10867 (10.2%) | < 0.01 |
| Past history of obesity | 2050 (31.71%) | 16482 (15.46%) | < 0.01 |
| Past history of preeclampsia | 525 (8.12%) | 2179 (2.04%) | < 0.01 |
| Past history of pregnancy related fatigue | 522 (8.08%) | 9311 (8.74%) | 0.08 |
| Past history of sexually transmitted disease | 345 (5.34%) | 5163 (4.84%) | 0.081 |
| Chronic or gestational hypertension | 1239 (19.17%) | 3389 (3.18%) | < 0.01 |
| Gestational diabetes | 817 (12.64%) | 9424 (8.84%) | < 0.01 |
| Anemia during pregnancy | 900 (13.92%) | 12490 (11.72%) | < 0.01 |
| Headaches during pregnancy | 1016 (15.72%) | 11514 (10.8%) | < 0.01 |
| Autoimmune disease | 635 (9.82%) | 4902 (4.6%) | < 0.01 |
| High risk pregnancy | 3255 (50.36%) | 43013 (40.36%) | < 0.01 |
| Hyperemesis gravidarum | 445 (6.88%) | 5960 (5.59%) | < 0.01 |
| Pregnancy related fatigue | 793 (12.27%) | 11426 (10.72%) | < 0.01 |
| Proteinuria 0-28 weeks | 195 (3.02%) | 87 (0.08%) | < 0.01 |
| Maximal AST 0-28 weeks, | 17.0 (17.0 - 17.0) | 17.0 (17.0 - 17.0) | < 0.01 |
| Maximal white blood count 0-28 weeks | 9.0 (8.48 - 9.86) | 9.0 (8.33 - 9.6) | < 0.01 |
| Maximal ALT 0-28 weeks | 15.0 (15.0 - 15.0) | 15.0 (15.0 - 15.0) | < 0.01 |
| Maximal serum calcium 0-28 weeks | 9.3 (9.3 - 9.3) | 9.3 (9.3 - 9.3) | < 0.01 |
| Maximal serum creatinine 0-28 weeks | 0.59 (0.59 - 0.59) | 0.59 (0.59 - 0.59) | < 0.01 |
| Maximal eosinophils 0-28 weeks | 1.1 (1.1 - 1.1) | 1.1 (1.1 - 1.1) | 0.019 |
| Maximal serum glucose 0-28 weeks | 87.0 (87.0 - 87.0) | 87.0 (87.0 - 87.0) | < 0.01 |
| Minimal hemoglobin 0-28 weeks | 11.6 (11.1 - 12.1) | 11.6 (11.2 - 12.1) | 0.24 |
| Maximal lymphocytes 0-28 weeks | 22.7 (22.7 - 22.7) | 22.7 (22.7 - 22.7) | 0.65 |
| Maximal platelets 0-28 weeks | 258.0 (244.0 - 306.0) | 258.0 (233.0 - 283.0) | < 0.01 |
| Minimal red blood cell count 0-28 weeks | 3.82 (3.69 - 4.1) | 3.82 (3.68 - 4.01) | < 0.01 |
| Antihypertensive medications 0-28 weeks | 167 (2.58%) | 117 (0.11%) | < 0.01 |

Median (IQR) for continuous variables; n (%) for categorical variables; p-values for continuous variables based on Kruskal-Wallis rank sum test; for categorical variables based on Fisher's exact or Chi-squared test.
Abbreviations: SBP, systolic blood pressure; DBP, diastolic blood pressure, BMI, body mass index, PT/INR, prothrombin time/international normalized ratio; AST, aspartate transferase; ALT, alanine transaminase; SGA, small for gestational age; IUGR, intrauterine growth restriction.

Table S9. Summary characteristics of Cohort 1 patient data at 32 weeks gestation.

| | **Preeclampsia (n=6464)** | **No preeclampsia (n=106581)** | **P-Value** |
|---|---|---|---|
| Maternal age, y | 33.03 (29.66 - 36.65) | 32.99 (30.03 - 35.9) | 0.12 |
| Self-reported race, White | 3825 (59.17%) | 71037 (66.65%) | < 0.01 |
| Self-reported race, Black | 1166 (18.04%) | 10283 (9.65%) | < 0.01 |
| Self-reported race, other | 1517 (23.47%) | 25798 (24.2%) | 0.24 |
| Self-reported ethnicity, Hispanic | 1263 (19.54%) | 16704 (15.67%) | < 0.01 |
| Self-reported ethnicity, non-Hispanic | 5201 (80.46%) | 89879 (84.33%) | < 0.01 |
| Private insurance | 5085 (78.67%) | 90488 (84.9%) | < 0.01 |
| Public insurance | 1851 (28.64%) | 22076 (20.71%) | < 0.01 |
| Alcohol use history | 1577 (24.4%) | 24164 (22.67%) | < 0.01 |
| Smoking history | 877 (13.57%) | 10403 (9.76%) | < 0.01 |
| Illicit drugs history | 317 (4.9%) | 3213 (3.01%) | < 0.01 |
| Gravidity | 2.0 (1.0 - 3.0) | 2.0 (1.0 - 3.0) | 0.61 |
| Parity | 1.0 (1.0 - 1.0) | 1.0 (1.0 - 1.0) | < 0.01 |
| In vitro fertilization | 396 (6.13%) | 4141 (3.89%) | < 0.01 |
| Nulliparous | 1183 (18.3%) | 13790 (12.94%) | < 0.01 |
| Interpregnancy interval, y | 2.3 (2.3 - 2.3) | 2.3 (2.3 - 2.3) | 0.93 |
| Multiple gestation | 479 (7.41%) | 2870 (2.69%) | < 0.01 |
| Maximal SBP 0-32 weeks, mmHg | 130.0 (122.0 - 142.0) | 122.0 (117.0 - 126.0) | < 0.01 |
| Maximal DBP 0-32 weeks, mmHg | 80.0 (75.0 - 88.0) | 75.0 (70.0 - 80.0) | < 0.01 |
| Maximal heart rate 0-32 weeks, bpm | 91.0 (91.0 - 96.0) | 91.0 (91.0 - 91.0) | < 0.01 |
| Maximal body mass index 0-32 weeks, ($kg/m^2$) | 29.3 (28.3 - 35.1) | 28.3 (26.3 - 30.2) | < 0.01 |
| Maximal weight 0-32 weeks, kg | 78.47 (74.57 - 93.9) | 75.75 (69.58 - 81.65) | < 0.01 |
| Family history of chronic hypertension | 2907 (44.97%) | 38881 (36.48%) | < 0.01 |
| Family history of heart disease | 2980 (46.1%) | 40548 (38.04%) | < 0.01 |
| Family history of preeclampsia | 144 (2.23%) | 2135 (2.0%) | 0.22 |
| Family history of diabetes | 329 (5.09%) | 4078 (3.83%) | < 0.01 |
| Family history of hyperlipidemia | 120 (1.86%) | 1571 (1.47%) | 0.015 |
| Family history of stroke | 157 (2.43%) | 1854 (1.74%) | < 0.01 |
| Past history of diabetes | 507 (7.84%) | 3036 (2.85%) | < 0.01 |
| Past history of cesarean delivery | 1458 (22.56%) | 19675 (18.46%) | < 0.01 |
| Past history of preterm birth | 510 (7.89%) | 4582 (4.3%) | < 0.01 |
| Past history of gynecologic surgery | 780 (12.07%) | 13528 (12.69%) | 0.17 |
| Past history of asthma | 949 (14.68%) | 11136 (10.45%) | < 0.01 |
| Past history of chronic hypertension | 1868 (28.9%) | 5725 (5.37%) | < 0.01 |
| Past history of gestational hypertension | 500 (7.74%) | 3451 (3.24%) | < 0.01 |
| Past history of high-risk pregnancy | 1166 (18.04%) | 18472 (17.33%) | 0.19 |
| Past history of hyperemesis gravidarum | 1037 (16.04%) | 17344 (16.27%) | 0.66 |
| Past history of migraine | 900 (13.92%) | 10867 (10.2%) | < 0.01 |
| Past history of obesity | 2050 (31.71%) | 16482 (15.46%) | < 0.01 |
| Past history of preeclampsia | 525 (8.12%) | 2179 (2.04%) | < 0.01 |
| Past history of pregnancy related fatigue | 522 (8.08%) | 9311 (8.74%) | 0.08 |
| Past history of sexually transmitted disease | 345 (5.34%) | 5163 (4.84%) | 0.081 |
| Chronic or gestational hypertension | 1534 (23.73%) | 4052 (3.8%) | < 0.01 |
| Gestational diabetes | 1087 (16.82%) | 12544 (11.77%) | < 0.01 |
| Anemia during pregnancy | 1027 (15.89%) | 14487 (13.59%) | < 0.01 |
| Headaches during pregnancy | 1040 (16.09%) | 11681 (10.96%) | < 0.01 |
| Autoimmune disease | 635 (9.82%) | 4902 (4.6%) | < 0.01 |
| High risk pregnancy | 3620 (56.0%) | 47635 (44.69%) | < 0.01 |
| Hyperemesis gravidarum | 536 (8.29%) | 7324 (6.87%) | < 0.01 |
| Pregnancy related fatigue | 1149 (17.78%) | 16390 (15.38%) | < 0.01 |
| Proteinuria 0-32 weeks | 305 (4.72%) | 130 (0.12%) | < 0.01 |
| Maximal AST 0-32 weeks | 17.0 (17.0 - 17.0) | 17.0 (17.0 - 17.0) | < 0.01 |
| Maximal uric acid 0-32 weeks | 3.6 (3.6 - 3.6) | 3.6 (3.6 - 3.6) | < 0.01 |
| Maximal white blood count 0-32 weeks | 9.1 (8.57 - 9.9) | 9.1 (8.45 - 9.67) | < 0.01 |
| Maximal ALT 0-32 weeks | 15.0 (15.0 - 15.0) | 15.0 (15.0 - 15.0) | < 0.01 |
| Maximal serum calcium 0-32 weeks | 9.3 (9.3 - 9.3) | 9.3 (9.3 - 9.3) | < 0.01 |
| Maximal serum creatinine 0-32 weeks | 0.59 (0.59 - 0.6) | 0.59 (0.59 - 0.59) | < 0.01 |
| Maximal eosinophils 0-32 weeks | 1.1 (1.1 - 1.2) | 1.1 (1.1 - 1.1) | 0.017 |
| Maximal serum glucose 0-32 weeks | 86.0 (86.0 - 89.0) | 86.0 (86.0 - 86.0) | < 0.01 |
| Minimal hemoglobin 0-32 weeks | 11.5 (10.9 - 11.9) | 11.5 (11.0 - 11.9) | 0.48 |
| Maximal lymphocytes 0-32 weeks | 22.6 (22.6 - 22.6) | 22.6 (22.6 - 22.6) | 0.74 |
| Maximal platelets 0-32 weeks | 261.5 (242.0 - 308.0) | 258.0 (231.0 - 285.0) | 0.025 |
| Minimal red blood cell count 0-32 weeks | 3.75 (3.62 - 4.01) | 3.75 (3.61 - 3.91) | < 0.01 |
| Antihypertensive medications 0-32 weeks | 263 (4.07%) | 185 (0.17%) | < 0.01 |

Median (IQR) for continuous variables; n (%) for categorical variables; p-values for continuous variables based on Kruskal-Wallis rank sum test; for categorical variables based on Fisher's exact or Chi-squared test.
Abbreviations: SBP, systolic blood pressure; DBP, diastolic blood pressure, BMI, body mass index, PT/INR, prothrombin time/international normalized ratio; AST, aspartate transferase; ALT, alanine transaminase; SGA, small for gestational age; IUGR, intrauterine growth restriction.

Table S10. Summary characteristics of Cohort 1 patient data at 36 weeks gestation.

| | **Preeclampsia (n=6464)** | **No preeclampsia (n=106581)** | **P-Value** |
|---|---|---|---|
| Maternal age, y | 33.03 (29.66 - 36.65) | 32.99 (30.03 - 35.9) | 0.12 |
| Self-reported race, White | 3825 (59.17%) | 71037 (66.65%) | < 0.01 |
| Self-reported race, Black | 1166 (18.04%) | 10283 (9.65%) | < 0.01 |
| Self-reported race, other | 1517 (23.47%) | 25798 (24.2%) | 0.24 |
| Self-reported ethnicity, Hispanic | 1263 (19.54%) | 16704 (15.67%) | < 0.01 |
| Self-reported ethnicity, non-Hispanic | 5201 (80.46%) | 89879 (84.33%) | < 0.01 |
| Private insurance | 5085 (78.67%) | 90488 (84.9%) | < 0.01 |
| Public insurance | 1851 (28.64%) | 22076 (20.71%) | < 0.01 |
| Alcohol use history | 1577 (24.4%) | 24164 (22.67%) | < 0.01 |
| Smoking history | 877 (13.57%) | 10403 (9.76%) | < 0.01 |
| Illicit drugs history | 317 (4.9%) | 3213 (3.01%) | < 0.01 |
| Gravidity | 2.0 (1.0 - 3.0) | 2.0 (1.0 - 3.0) | 0.61 |
| Parity | 1.0 (1.0 - 1.0) | 1.0 (1.0 - 1.0) | < 0.01 |
| In vitro fertilization | 396 (6.13%) | 4141 (3.89%) | < 0.01 |
| Nulliparous | 1183 (18.3%) | 13790 (12.94%) | < 0.01 |
| Interpregnancy interval, y | 2.3 (2.3 - 2.3) | 2.3 (2.3 - 2.3) | 0.93 |
| Multiple gestation | 479 (7.41%) | 2870 (2.69%) | < 0.01 |
| Maximal SBP 0-36 weeks, mmHg | 136.0 (124.0 - 150.0) | 124.0 (118.0 - 128.0) | < 0.01 |
| Maximal DBP 0-36 weeks, mmHg | 84.0 (78.0 - 93.0) | 77.0 (71.0 - 80.0) | < 0.01 |
| Maximal heart rate 0-36 weeks, bpm | 93.0 (93.0 - 100.0) | 93.0 (93.0 - 93.0) | < 0.01 |
| Maximal body mass index 0-36 weeks, ($kg/m^2$) | 30.8 (29.2 - 36.3) | 29.2 (27.0 - 31.2) | < 0.01 |
| Maximal weight 0-36 weeks, kg | 82.1 (76.66 - 97.52) | 78.02 (71.3 - 84.37) | < 0.01 |
| Family history of chronic hypertension | 2907 (44.97%) | 38881 (36.48%) | < 0.01 |
| Family history of heart disease | 2980 (46.1%) | 40548 (38.04%) | < 0.01 |
| Family history of preeclampsia | 144 (2.23%) | 2135 (2.0%) | 0.22 |
| Family history of diabetes | 329 (5.09%) | 4078 (3.83%) | < 0.01 |
| Family history of hyperlipidemia | 120 (1.86%) | 1571 (1.47%) | 0.015 |
| Family history of stroke | 157 (2.43%) | 1854 (1.74%) | < 0.01 |
| Past history of diabetes | 507 (7.84%) | 3036 (2.85%) | < 0.01 |
| Past history of cesarean delivery | 1458 (22.56%) | 19675 (18.46%) | < 0.01 |
| Past history of preterm birth | 510 (7.89%) | 4582 (4.3%) | < 0.01 |
| Past history of gynecologic surgery | 780 (12.07%) | 13528 (12.69%) | 0.17 |
| Past history of asthma | 949 (14.68%) | 11136 (10.45%) | < 0.01 |
| Past history of chronic hypertension | 1868 (28.9%) | 5725 (5.37%) | < 0.01 |
| Past history of gestational hypertension | 500 (7.74%) | 3451 (3.24%) | < 0.01 |
| Past history of high-risk pregnancy | 1166 (18.04%) | 18472 (17.33%) | 0.19 |
| Past history of hyperemesis gravidarum | 1037 (16.04%) | 17344 (16.27%) | 0.66 |
| Past history of migraine | 900 (13.92%) | 10867 (10.2%) | < 0.01 |
| Past history of obesity | 2050 (31.71%) | 16482 (15.46%) | < 0.01 |
| Past history of preeclampsia | 525 (8.12%) | 2179 (2.04%) | < 0.01 |
| Past history of pregnancy related fatigue | 522 (8.08%) | 9311 (8.74%) | 0.08 |
| Past history of sexually transmitted disease | 345 (5.34%) | 5163 (4.84%) | 0.081 |
| Chronic or gestational hypertension | 2115 (32.72%) | 5558 (5.21%) | < 0.01 |
| Gestational diabetes | 1160 (17.95%) | 13122 (12.31%) | < 0.01 |
| Anemia during pregnancy | 1121 (17.34%) | 15649 (14.68%) | < 0.01 |
| Headaches during pregnancy | 1092 (16.89%) | 11925 (11.19%) | < 0.01 |
| Autoimmune disease | 635 (9.82%) | 4902 (4.6%) | < 0.01 |
| High risk pregnancy | 3813 (58.99%) | 50232 (47.13%) | < 0.01 |
| Hyperemesis gravidarum | 564 (8.73%) | 7964 (7.47%) | < 0.01 |
| Pregnancy related fatigue | 1271 (19.66%) | 18368 (17.23%) | < 0.01 |
| Proteinuria 0-36 weeks | 589 (9.11%) | 211 (0.2%) | < 0.01 |
| Maximal AST 0-36 weeks | 18.0 (18.0 - 21.0) | 18.0 (18.0 - 18.0) | < 0.01 |
| Maximal lactate dehydrogenase 0-36 weeks | 181.0 (181.0 - 181.0) | 181.0 (181.0 - 181.0) | < 0.01 |
| Maximal uric acid 0-36 weeks | 4.0 (4.0 - 4.4) | 4.0 (4.0 - 4.0) | < 0.01 |
| Maximal white blood count 0-36 weeks | 9.16 (8.61 - 9.9) | 9.14 (8.47 - 9.7) | < 0.01 |
| Maximal ALT 0-36 weeks | 15.0 (15.0 - 17.0) | 15.0 (15.0 - 15.0) | < 0.01 |
| Maximal serum calcium 0-36 weeks | 9.3 (9.3 - 9.3) | 9.3 (9.3 - 9.3) | < 0.01 |
| Maximal serum creatinine 0-36 weeks | 0.59 (0.59 - 0.65) | 0.59 (0.59 - 0.59) | < 0.01 |
| Maximal eosinophils 0-36 weeks | 1.1 (1.1 - 1.2) | 1.1 (1.1 - 1.1) | 0.11 |
| Maximal serum glucose 0-36 weeks | 87.0 (87.0 - 92.0) | 87.0 (87.0 - 87.0) | < 0.01 |
| Minimal hemoglobin 0-36 weeks | 11.4 (10.8 - 11.9) | 11.4 (11.0 - 11.9) | < 0.01 |
| Maximal lymphocytes 0-36 weeks | 22.5 (22.5 - 22.5) | 22.5 (22.5 - 22.5) | 0.75 |
| Maximal platelets 0-36 weeks | 264.0 (239.0 - 310.0) | 257.0 (230.0 - 286.0) | 0.045 |
| Minimal red blood cell count 0-36 weeks | 3.75 (3.6 - 4.0) | 3.75 (3.6 - 3.91) | < 0.01 |
| Antihypertensive medications 0-36 weeks | 518 (8.01%) | 1128 (1.06%) | < 0.01 |

Median (IQR) for continuous variables; n (%) for categorical variables; p-values for continuous variables based on Kruskal-Wallis rank sum test; for categorical variables based on Fisher's exact or Chi-squared test.
Abbreviations: SBP, systolic blood pressure; DBP, diastolic blood pressure, BMI, body mass index, PT/INR, prothrombin time/international normalized ratio; AST, aspartate transferase; ALT, alanine transaminase; SGA, small for gestational age; IUGR, intrauterine growth restriction.

Table S11. Summary characteristics of Cohort 1 patient data at 39 weeks gestation.

| | **Preeclampsia (n=6464)** | **No preeclampsia (n=106581)** | **P-Value** |
|---|---|---|---|
| Maternal age, y | 33.03 (29.66 - 36.65) | 32.99 (30.03 - 35.9) | 0.12 |
| Self-reported race, White | 3825 (59.17%) | 71037 (66.65%) | < 0.01 |
| Self-reported race, Black | 1166 (18.04%) | 10283 (9.65%) | < 0.01 |
| Self-reported race, other | 1517 (23.47%) | 25798 (24.2%) | 0.24 |
| Self-reported ethnicity, Hispanic | 1263 (19.54%) | 16704 (15.67%) | < 0.01 |
| Self-reported ethnicity, non-Hispanic | 5201 (80.46%) | 89879 (84.33%) | < 0.01 |
| Private insurance | 5085 (78.67%) | 90488 (84.9%) | < 0.01 |
| Public insurance | 1851 (28.64%) | 22076 (20.71%) | < 0.01 |
| Alcohol use history | 1577 (24.4%) | 24164 (22.67%) | < 0.01 |
| Smoking history | 877 (13.57%) | 10403 (9.76%) | < 0.01 |
| Illicit drugs history | 317 (4.9%) | 3213 (3.01%) | < 0.01 |
| Gravidity | 2.0 (1.0 - 3.0) | 2.0 (1.0 - 3.0) | 0.61 |
| Parity | 1.0 (1.0 - 1.0) | 1.0 (1.0 - 1.0) | < 0.01 |
| In vitro fertilization | 396 (6.13%) | 4141 (3.89%) | < 0.01 |
| Nulliparous | 1183 (18.3%) | 13790 (12.94%) | < 0.01 |
| Interpregnancy interval, y | 2.3 (2.3 - 2.3) | 2.3 (2.3 - 2.3) | 0.93 |
| Multiple gestation | 479 (7.41%) | 2870 (2.69%) | < 0.01 |
| Maximal SBP 0-39 weeks, mmHg | 144.0 (130.0 - 158.0) | 127.0 (120.0 - 132.0) | < 0.01 |
| Maximal DBP 0-39 weeks, mmHg | 90.0 (82.0 - 98.0) | 80.0 (74.0 - 82.0) | < 0.01 |
| Maximal heart rate 0-39 weeks, bpm | 94.0 (91.0 - 104.0) | 94.0 (92.0 - 96.0) | < 0.01 |
| Maximal body mass index 0-39 weeks, (kg/m$^2$) | 31.83 (29.8 - 37.2) | 29.9 (27.5 - 32.2) | < 0.01 |
| Maximal weight 0-39 weeks, kg | 84.37 (77.56 - 99.34) | 79.83 (72.67 - 86.91) | < 0.01 |
| Family history of chronic hypertension | 2907 (44.97%) | 38881 (36.48%) | < 0.01 |
| Family history of heart disease | 2980 (46.1%) | 40548 (38.04%) | < 0.01 |
| Family history of preeclampsia | 144 (2.23%) | 2135 (2.0%) | 0.22 |
| Family history of diabetes | 329 (5.09%) | 4078 (3.83%) | < 0.01 |
| Family history of hyperlipidemia | 120 (1.86%) | 1571 (1.47%) | 0.015 |
| Family history of stroke | 157 (2.43%) | 1854 (1.74%) | < 0.01 |
| Past history of diabetes | 507 (7.84%) | 3036 (2.85%) | < 0.01 |
| Past history of cesarean delivery | 1458 (22.56%) | 19675 (18.46%) | < 0.01 |
| Past history of preterm birth | 510 (7.89%) | 4582 (4.3%) | < 0.01 |
| Past history of gynecologic surgery | 780 (12.07%) | 13528 (12.69%) | 0.17 |
| Past history of asthma | 949 (14.68%) | 11136 (10.45%) | < 0.01 |
| Past history of chronic hypertension | 1868 (28.9%) | 5725 (5.37%) | < 0.01 |
| Past history of gestational hypertension | 500 (7.74%) | 3451 (3.24%) | < 0.01 |
| Past history of high-risk pregnancy | 1166 (18.04%) | 18472 (17.33%) | 0.19 |
| Past history of hyperemesis gravidarum | 1037 (16.04%) | 17344 (16.27%) | 0.66 |
| Past history of migraine | 900 (13.92%) | 10867 (10.2%) | < 0.01 |
| Past history of obesity | 2050 (31.71%) | 16482 (15.46%) | < 0.01 |
| Past history of preeclampsia | 525 (8.12%) | 2179 (2.04%) | < 0.01 |
| Past history of pregnancy related fatigue | 522 (8.08%) | 9311 (8.74%) | 0.08 |
| Past history of sexually transmitted disease | 345 (5.34%) | 5163 (4.84%) | 0.081 |
| Chronic or gestational hypertension | 2660 (41.15%) | 7893 (7.41%) | < 0.01 |
| Gestational diabetes | 1181 (18.27%) | 13379 (12.55%) | < 0.01 |
| Anemia during pregnancy | 1179 (18.24%) | 16301 (15.29%) | < 0.01 |
| Headaches during pregnancy | 1119 (17.31%) | 12163 (11.41%) | < 0.01 |
| Autoimmune disease | 635 (9.82%) | 4902 (4.6%) | < 0.01 |
| High risk pregnancy | 3938 (60.92%) | 52532 (49.29%) | < 0.01 |
| Hyperemesis gravidarum | 578 (8.94%) | 8339 (7.82%) | < 0.01 |
| Pregnancy related fatigue | 1323 (20.47%) | 19439 (18.24%) | < 0.01 |
| Gestational diabetes | 1181 (18.27%) | 13379 (12.55%) | < 0.01 |
| Oligohydramnios | 144 (2.23%) | 1805 (1.69%) | < 0.01 |
| Proteinuria 0-39 weeks | 1130 (17.48%) | 339 (0.32%) | < 0.01 |
| Maximal AST 0-39 weeks | 18.0 (18.0 - 24.0) | 18.0 (18.0 - 18.0) | < 0.01 |
| Maximal lactate dehydrogenase 0-39 weeks | 186.0 (186.0 - 189.0) | 186.0 (186.0 - 186.0) | < 0.01 |
| Maximal uric acid 0-39 weeks | 4.4 (4.4 - 5.3) | 4.4 (4.4 - 4.4) | < 0.01 |
| Maximal white blood count 0-39 weeks | 9.28 (8.63 - 9.9) | 9.2 (8.51 - 9.75) | < 0.01 |
| Maximal ALT 0-39 weeks | 14.0 (13.0 - 20.0) | 14.0 (14.0 - 14.0) | < 0.01 |
| Maximal serum calcium 0-39 weeks | 9.3 (9.3 - 9.3) | 9.3 (9.3 - 9.3) | < 0.01 |
| Maximal serum creatinine 0-39 weeks | 0.6 (0.6 - 0.7) | 0.6 (0.6 - 0.6) | < 0.01 |
| Maximal eosinophils 0-39 weeks | 1.1 (1.1 - 1.2) | 1.1 (1.1 - 1.1) | 0.11 |
| Maximal serum glucose 0-39 weeks | 87.0 (86.0 - 93.0) | 87.0 (87.0 - 87.0) | < 0.01 |
| Minimal hemoglobin 0-39 weeks | 11.4 (10.7 - 11.82) | 11.4 (10.9 - 11.9) | < 0.01 |
| Maximal lymphocytes 0-39 weeks | 22.5 (22.5 - 22.5) | 22.5 (22.5 - 22.5) | 0.65 |
| Maximal platelets 0-39 weeks | 265.0 (235.0 - 311.0) | 257.0 (229.0 - 287.0) | 0.048 |

| | | | |
|---|---|---|---|
| Minimal red blood cell count 0-39 weeks | 3.75 (3.58 - 4.01) | 3.75 (3.6 - 3.92) | < 0.01 |
| Antihypertensive medications 0-39 weeks | 760 (11.76%) | 1587 (1.49%) | < 0.01 |

Median (IQR) for continuous variables; n (%) for categorical variables; p-values for continuous variables based on Kruskal-Wallis rank sum test; for categorical variables based on Fisher's exact or Chi-squared test.
Abbreviations: SBP, systolic blood pressure; DBP, diastolic blood pressure, BMI, body mass index, PT/INR, prothrombin time/international normalized ratio; AST, aspartate transferase; ALT, alanine transaminase; SGA, small for gestational age; IUGR, intrauterine growth restriction.

Table S12. Summary characteristics of Cohort 1 patient data on admission.

| | **Preeclampsia (n=6464)** | **No preeclampsia (n=106581)** | **P-Value** |
|---|---|---|---|
| Maternal age, y | 33.03 (29.66 - 36.65) | 32.99 (30.03 - 35.9) | 0.12 |
| Self-reported race, White | 3825 (59.17%) | 71037 (66.65%) | < 0.01 |
| Self-reported race, Black | 1166 (18.04%) | 10283 (9.65%) | < 0.01 |
| Self-reported race, other | 1517 (23.47%) | 25798 (24.2%) | 0.24 |
| Self-reported ethnicity, Hispanic | 1263 (19.54%) | 16704 (15.67%) | < 0.01 |
| Self-reported ethnicity, non-Hispanic | 5201 (80.46%) | 89879 (84.33%) | < 0.01 |
| Private insurance | 5085 (78.67%) | 90488 (84.9%) | < 0.01 |
| Public insurance | 1851 (28.64%) | 22076 (20.71%) | < 0.01 |
| Alcohol use history | 1577 (24.4%) | 24164 (22.67%) | < 0.01 |
| Smoking history | 877 (13.57%) | 10403 (9.76%) | < 0.01 |
| Illicit drugs history | 317 (4.9%) | 3213 (3.01%) | < 0.01 |
| Gravidity | 2.0 (1.0 - 3.0) | 2.0 (1.0 - 3.0) | 0.61 |
| Parity | 1.0 (1.0 - 1.0) | 1.0 (1.0 - 1.0) | < 0.01 |
| In vitro fertilization | 396 (6.13%) | 4141 (3.89%) | < 0.01 |
| Nulliparous | 1183 (18.3%) | 13790 (12.94%) | < 0.01 |
| Interpregnancy interval, y | 2.3 (2.3 - 2.3) | 2.3 (2.3 - 2.3) | 0.93 |
| Multiple gestation | 479 (7.41%) | 2870 (2.69%) | < 0.01 |
| Maximal SBP 0 weeks-admission, mmHg | 148.0 (139.0 - 160.0) | 130.0 (122.0 - 137.0) | < 0.01 |
| Maximal DBP 0 weeks-admission, mmHg | 92.0 (85.0 - 99.0) | 80.0 (75.0 - 85.0) | < 0.01 |
| Maximal heart rate 0 weeks-admission, bpm | 95.0 (90.0 - 106.0) | 95.0 (89.0 - 100.0) | < 0.01 |
| Maximal body mass index 0 weeks-admission, (kg/m$^2$) | 32.4 (29.7 - 37.5) | 30.1 (27.5 - 32.7) | < 0.01 |
| Maximal weight 0 weeks-admission, kg | 85.28 (77.56 - 100.12) | 80.29 (72.67 - 88.0) | < 0.01 |
| Family history of chronic hypertension | 2907 (44.97%) | 38881 (36.48%) | < 0.01 |
| Family history of heart disease | 2980 (46.1%) | 40548 (38.04%) | < 0.01 |
| Family history of preeclampsia | 144 (2.23%) | 2135 (2.0%) | 0.22 |
| Family history of diabetes | 329 (5.09%) | 4078 (3.83%) | < 0.01 |
| Family history of hyperlipidemia | 120 (1.86%) | 1571 (1.47%) | 0.015 |
| Family history of stroke | 157 (2.43%) | 1854 (1.74%) | < 0.01 |
| Past history of diabetes | 507 (7.84%) | 3036 (2.85%) | < 0.01 |
| Past history of cesarean delivery | 1458 (22.56%) | 19675 (18.46%) | < 0.01 |
| Past history of preterm birth | 510 (7.89%) | 4582 (4.3%) | < 0.01 |
| Past history of gynecologic surgery | 780 (12.07%) | 13528 (12.69%) | 0.17 |
| Past history of asthma | 949 (14.68%) | 11136 (10.45%) | < 0.01 |
| Past history of chronic hypertension | 1868 (28.9%) | 5725 (5.37%) | < 0.01 |
| Past history of gestational hypertension | 500 (7.74%) | 3451 (3.24%) | < 0.01 |
| Past history of high-risk pregnancy | 1166 (18.04%) | 18472 (17.33%) | 0.19 |
| Past history of hyperemesis gravidarum | 1037 (16.04%) | 17344 (16.27%) | 0.66 |
| Past history of migraine | 900 (13.92%) | 10867 (10.2%) | < 0.01 |
| Past history of obesity | 2050 (31.71%) | 16482 (15.46%) | < 0.01 |
| Past history of preeclampsia | 525 (8.12%) | 2179 (2.04%) | < 0.01 |
| Past history of pregnancy related fatigue | 522 (8.08%) | 9311 (8.74%) | 0.08 |
| Past history of sexually transmitted disease | 345 (5.34%) | 5163 (4.84%) | 0.081 |
| Chronic or gestational hypertension | 2780 (43.01%) | 8597 (8.07%) | < 0.01 |
| Gestational diabetes | 1185 (18.33%) | 13440 (12.61%) | < 0.01 |
| Anemia during pregnancy | 1205 (18.64%) | 16841 (15.8%) | < 0.01 |
| Headaches during pregnancy | 1121 (17.34%) | 12257 (11.5%) | < 0.01 |
| Autoimmune disease | 635 (9.82%) | 4902 (4.6%) | < 0.01 |
| High risk pregnancy | 3960 (61.26%) | 53235 (49.95%) | < 0.01 |
| Hyperemesis gravidarum | 580 (8.97%) | 8407 (7.89%) | < 0.01 |
| Pregnancy related fatigue | 1328 (20.54%) | 19687 (18.47%) | < 0.01 |
| Gestational diabetes | 1185 (18.33%) | 13440 (12.61%) | < 0.01 |
| Oligohydramnios | 157 (2.43%) | 2157 (2.02%) | 0.027 |
| Proteinuria 0 weeks-admission | 1430 (22.12%) | 419 (0.39%) | < 0.01 |
| Maximal AST 0 weeks-admission | 19.0 (18.0 - 25.0) | 19.0 (19.0 - 19.0) | < 0.01 |
| Maximal lactate dehydrogenase 0 weeks-admission | 187.0 (187.0 - 200.0) | 187.0 (187.0 - 187.0) | < 0.01 |
| Maximal uric acid 0 weeks-admission | 4.5 (4.5 - 5.5) | 4.5 (4.5 - 4.5) | < 0.01 |
| Maximal white blood count 0 weeks-admission | 9.35 (8.66 - 9.92) | 9.29 (8.55 - 9.81) | < 0.01 |
| Maximal ALT 0 weeks-admission | 14.0 (12.0 - 21.0) | 14.0 (14.0 - 14.0) | < 0.01 |
| Maximal serum calcium 0 weeks-admission | 9.3 (9.3 - 9.3) | 9.3 (9.3 - 9.3) | < 0.01 |
| Maximal serum creatinine 0 weeks-admission | 0.61 (0.6 - 0.72) | 0.6 (0.6 - 0.6) | < 0.01 |
| Maximal eosinophils 0 weeks-admission | 1.1 (1.1 - 1.2) | 1.1 (1.1 - 1.1) | 0.1 |
| Maximal serum glucose 0 weeks-admission | 87.0 (86.0 - 94.0) | 87.0 (87.0 - 87.0) | < 0.01 |
| Minimal hemoglobin 0 weeks-admission | 11.3 (10.7 - 11.9) | 11.4 (10.9 - 11.9) | < 0.01 |
| Maximal lymphocytes 0 weeks-admission | 22.5 (22.5 - 22.5) | 22.5 (22.5 - 22.5) | 0.63 |
| Maximal platelets 0 weeks-admission | 266.0 (233.0 - 311.0) | 255.0 (225.0 - 288.0) | 0.027 |

| | | | |
|---|---|---|---|
| Minimal red blood cell count 0 weeks-admission | 3.77 (3.57 - 4.03) | 3.77 (3.59 - 3.95) | < 0.01 |
| Antihypertensive medications 0 weeks-admission | 821 (12.7%) | 1790 (1.68%) | < 0.01 |

Median (IQR) for continuous variables; n (%) for categorical variables; p-values for continuous variables based on Kruskal-Wallis rank sum test; for categorical variables based on Fisher's exact or Chi-squared test.
Abbreviations: SBP, systolic blood pressure; DBP, diastolic blood pressure, BMI, body mass index, PT/INR, prothrombin time/international normalized ratio; AST, aspartate transferase; ALT, alanine transaminase; SGA, small for gestational age; IUGR, intrauterine growth restriction.

Table S13. Predictive performance metrics of all models in the test dataset from Cohort 1 by gestational age of model creation.

| | | AUC | specificity | sensitivity | PPV | NPV | accuracy | F1 |
|---|---|---|---|---|---|---|---|---|
| XGBoost | 14weeks | 0.79 | 0.78 | 0.62 | 0.15 | 0.97 | 0.77 | 0.24 |
| | 20weeks | 0.79 | 0.79 | 0.64 | 0.15 | 0.97 | 0.78 | 0.25 |
| | 24weeks | 0.78 | 0.79 | 0.62 | 0.15 | 0.97 | 0.78 | 0.24 |
| | 28weeks | 0.78 | 0.79 | 0.63 | 0.15 | 0.97 | 0.78 | 0.25 |
| | 32weeks | 0.81 | 0.8 | 0.67 | 0.17 | 0.98 | 0.79 | 0.27 |
| | 36weeks | 0.84 | 0.84 | 0.68 | 0.2 | 0.98 | 0.83 | 0.31 |
| | 39weeks | 0.89 | 0.87 | 0.75 | 0.25 | 0.98 | 0.86 | 0.38 |
| | admission | 0.91 | 0.88 | 0.79 | 0.29 | 0.99 | 0.88 | 0.42 |
| Neural Network | 14weeks | 0.77 | 1 | 0 | 1 | 0.94 | 0.94 | 0 |
| | 20weeks | 0.77 | 0.99 | 0.1 | 0.53 | 0.95 | 0.93 | 0.17 |
| | 24weeks | 0.53 | 0 | 1 | 0.06 | 0.95 | 0.06 | 0.11 |
| | 28weeks | 0.77 | 1 | 0.04 | 0.73 | 0.95 | 0.94 | 0.08 |
| | 32weeks | 0.79 | 1 | 0.1 | 0.69 | 0.95 | 0.94 | 0.17 |
| | 36weeks | 0.82 | 1 | 0.16 | 0.68 | 0.95 | 0.93 | 0.26 |
| | 39weeks | 0.86 | 1 | 0.22 | 0.74 | 0.95 | 0.93 | 0.34 |
| | admission | 0.9 | 1 | 0.27 | 0.77 | 0.96 | 0.93 | 0.4 |
| Elastic Net | 14weeks | 0.77 | 1 | 0.02 | 0.84 | 0.94 | 0.94 | 0.04 |
| | 20weeks | 0.77 | 1 | 0 | 1 | 0.94 | 0.94 | 0 |
| | 24weeks | 0.78 | 1 | 0.02 | 0.67 | 0.94 | 0.94 | 0.04 |
| | 28weeks | 0.79 | 1 | 0.04 | 0.72 | 0.94 | 0.94 | 0.07 |
| | 32weeks | 0.8 | 1 | 0.07 | 0.77 | 0.95 | 0.95 | 0.13 |
| | 36weeks | 0.82 | 1 | 0.11 | 0.76 | 0.95 | 0.95 | 0.2 |
| | 39weeks | 0.86 | 1 | 0.18 | 0.76 | 0.95 | 0.95 | 0.29 |
| | admission | 0.89 | 1 | 0.23 | 0.77 | 0.96 | 0.95 | 0.35 |
| Random Forest | 14weeks | 0.76 | 1 | 0.06 | 0.64 | 0.95 | 0.94 | 0.11 |
| | 20weeks | 0.77 | 1 | 0.06 | 0.7 | 0.95 | 0.94 | 0.11 |
| | 24weeks | 0.77 | 1 | 0.06 | 0.65 | 0.95 | 0.94 | 0.12 |
| | 28weeks | 0.78 | 1 | 0.08 | 0.68 | 0.95 | 0.95 | 0.15 |
| | 32weeks | 0.79 | 1 | 0.11 | 0.77 | 0.95 | 0.95 | 0.18 |
| | 36weeks | 0.81 | 1 | 0.18 | 0.75 | 0.95 | 0.95 | 0.29 |
| | 39weeks | 0.86 | 1 | 0.27 | 0.82 | 0.96 | 0.95 | 0.41 |
| | admission | 0.88 | 1 | 0.31 | 0.83 | 0.96 | 0.96 | 0.45 |
| Linear Regression | 14weeks | 0.77 | 1 | 0.02 | 0.67 | 0.94 | 0.94 | 0.04 |
| | 20weeks | 0.78 | 1 | 0.03 | 0.73 | 0.94 | 0.94 | 0.06 |
| | 24weeks | 0.78 | 1 | 0.03 | 0.64 | 0.94 | 0.94 | 0.06 |
| | 28weeks | 0.78 | 1 | 0.04 | 0.69 | 0.95 | 0.94 | 0.08 |
| | 32weeks | 0.79 | 1 | 0.07 | 0.75 | 0.95 | 0.95 | 0.13 |
| | 36weeks | 0.82 | 1 | 0.13 | 0.75 | 0.95 | 0.95 | 0.23 |
| | 39weeks | 0.86 | 1 | 0.2 | 0.77 | 0.95 | 0.95 | 0.32 |
| | admission | 0.89 | 1 | 0.24 | 0.78 | 0.96 | 0.95 | 0.37 |

Abbreviations: AUC, area under the curve, PPV, positive predictive value, NPV, negative predictive value.

### Supplementary Results

#### External validation of all predictive models

We further investigated the performance of our models using data from two community hospitals that were included in Cohort 2, 6.4% of all data (N=7,705); those were not used for model development and served to externally validate the results after all models were created and internally validated. The comparison of patient characteristics between Cohort 1 and Cohort 2 is shown in appendix pp 8-9. Cohort 2 had a similar incidence of preeclampsia as Cohort 1, 5.7% (N=421), but a larger proportion of patients with missing data, especially in the early gestational weeks.

We determined the generalizability of all models using Cohort 2 as an external validation dataset. While these two community hospitals are part of our healthcare system, each hospital is independent in its clinical management, and the physicians in each of the hospitals included in Cohort 2 practice only in that hospital. As a result, this Cohort 2 is sufficiently different from Cohort 1. In early pregnancy, at 14 weeks, the AUC range was 0.64-0.66. As the gestational age advanced, the predictive power of the models increased progressively, AUC range of 0.66-0.90 (appendix p 26).

Table S14. Predictive performance metrics of all models in the external validation dataset from Cohort 2 by gestational age of model creation.

| | Gestational age | AUC | Specificity | Sensitivity | PPV | NPV | accuracy | F1 |
|---|---|---|---|---|---|---|---|---|
| XGBoost | 14weeks | 0.66 | 0.86 | 0.33 | 0.13 | 0.95 | 0.83 | 0.18 |
| | 20weeks | 0.66 | 0.85 | 0.35 | 0.12 | 0.96 | 0.82 | 0.18 |
| | 24weeks | 0.67 | 0.85 | 0.37 | 0.13 | 0.96 | 0.82 | 0.19 |
| | 28weeks | 0.69 | 0.84 | 0.4 | 0.13 | 0.96 | 0.81 | 0.2 |
| | 32weeks | 0.71 | 0.83 | 0.44 | 0.14 | 0.96 | 0.81 | 0.21 |
| | 36weeks | 0.76 | 0.85 | 0.49 | 0.17 | 0.96 | 0.83 | 0.25 |
| | 39weeks | 0.86 | 0.88 | 0.66 | 0.25 | 0.98 | 0.86 | 0.36 |
| | admission | 0.9 | 0.88 | 0.75 | 0.28 | 0.98 | 0.87 | 0.41 |
| Neural Network | 14weeks | 0.64 | 1 | 0 | 0 | 0.94 | 0.94 | 0 |
| | 20weeks | 0.65 | 1 | 0.01 | 0.5 | 0.94 | 0.94 | 0.01 |
| | 24weeks | 0.66 | 1 | 0.02 | 0.41 | 0.94 | 0.94 | 0.03 |
| | 28weeks | 0.66 | 1 | 0.02 | 0.39 | 0.94 | 0.94 | 0.03 |
| | 32weeks | 0.69 | 1 | 0.05 | 0.48 | 0.94 | 0.94 | 0.09 |
| | 36weeks | 0.73 | 1 | 0.07 | 0.69 | 0.95 | 0.94 | 0.13 |
| | 39weeks | 0.84 | 1 | 0.02 | 1 | 0.94 | 0.94 | 0.04 |
| | admission | 0.89 | 1 | 0.18 | 0.86 | 0.95 | 0.93 | 0.3 |
| Elastic Net | 14weeks | 0.64 | 1 | 0 | 0 | 0.94 | 0.94 | 0 |
| | 20weeks | 0.64 | 1 | 0 | 0 | 0.94 | 0.94 | 0 |
| | 24weeks | 0.66 | 1 | 0 | 0.5 | 0.94 | 0.94 | 0 |
| | 28weeks | 0.68 | 1 | 0 | 0.4 | 0.94 | 0.94 | 0.01 |
| | 32weeks | 0.69 | 1 | 0 | 0.4 | 0.94 | 0.94 | 0.01 |
| | 36weeks | 0.74 | 1 | 0.05 | 0.75 | 0.94 | 0.94 | 0.09 |
| | 39weeks | 0.83 | 1 | 0.18 | 0.74 | 0.95 | 0.95 | 0.29 |
| | admission | 0.87 | 0.99 | 0.26 | 0.73 | 0.96 | 0.95 | 0.39 |
| Random Forest | 14weeks | 0.64 | 1 | 0 | 0.33 | 0.94 | 0.94 | 0.01 |
| | 20weeks | 0.64 | 1 | 0 | 0.25 | 0.94 | 0.94 | 0 |
| | 24weeks | 0.66 | 1 | 0.01 | 0.38 | 0.94 | 0.94 | 0.01 |
| | 28weeks | 0.68 | 1 | 0.01 | 0.67 | 0.94 | 0.94 | 0.03 |
| | 32weeks | 0.68 | 1 | 0.01 | 0.5 | 0.94 | 0.94 | 0.01 |
| | 36weeks | 0.72 | 1 | 0.05 | 0.9 | 0.94 | 0.94 | 0.09 |
| | 39weeks | 0.83 | 1 | 0.18 | 0.83 | 0.95 | 0.95 | 0.3 |
| | admission | 0.88 | 1 | 0.26 | 0.84 | 0.96 | 0.95 | 0.4 |
| Linear Regression | 14weeks | 0.64 | 1 | 0 | 0.17 | 0.94 | 0.94 | 0 |
| | 20weeks | 0.65 | 1 | 0 | 0.17 | 0.94 | 0.94 | 0 |
| | 24weeks | 0.66 | 1 | 0 | 0.17 | 0.94 | 0.94 | 0 |
| | 28weeks | 0.69 | 1 | 0 | 0.22 | 0.94 | 0.94 | 0.01 |
| | 32weeks | 0.69 | 1 | 0 | 0.2 | 0.94 | 0.94 | 0.01 |
| | 36weeks | 0.74 | 1 | 0.05 | 0.77 | 0.95 | 0.94 | 0.1 |
| | 39weeks | 0.83 | 1 | 0.18 | 0.73 | 0.95 | 0.95 | 0.29 |
| | admission | 0.87 | 0.99 | 0.27 | 0.73 | 0.96 | 0.95 | 0.39 |

Abbreviations: AUC, area under the curve, PPV, positive predictive value, NPV, negative predictive value.

**Performance based on racial and sociodemographic factors**

As our patient population was multi-racial and multi-ethnic, we explored the contribution of race and ethnicity to the model predictions. We selected the xgboost model for all experiments due to its high performance in our datasets. Initially, using all variables, including self-reported race and ethnicity, the model had an AUC of 0.91; the Shapley values demonstrated that Black race was the seventh most significant predictive feature at 14 weeks and remained in the top 20 features throughout pregnancy. Moreover, when evaluated in patients with self-reported Black race in the test dataset, the model had an AUC of 0.88; in contrast, the model had an AUC of 0.91 in patients with self-reported White race. We found that these differences in performance are due to a larger proportion of missing data in patients of Black race. When we evaluated the model performance in the subset of patients of Black race who had a similar rate of missingness to the rest of the cohort data, the performance of the model slightly improved, AUC of 0.90 (appendix p 28). Next, when we excluded race and ethnicity, the performance of the model slightly deteriorated, AUC of 0.90. As the model had the best performance when race and ethnicity were included, AUC of 0.91, we did all subsequent analyses using all variables available, including race and ethnicity.

Fig. S1 Including patients with complete data improves the performance of the xgboost model at preadmission in patients of different races. Initially, the model had AUC of 0.91 in all and White patients, but only 0.88 in Black patients. After removing 318 patients with more than 15% missing data, the AUC in Black patients increased to 0.90.

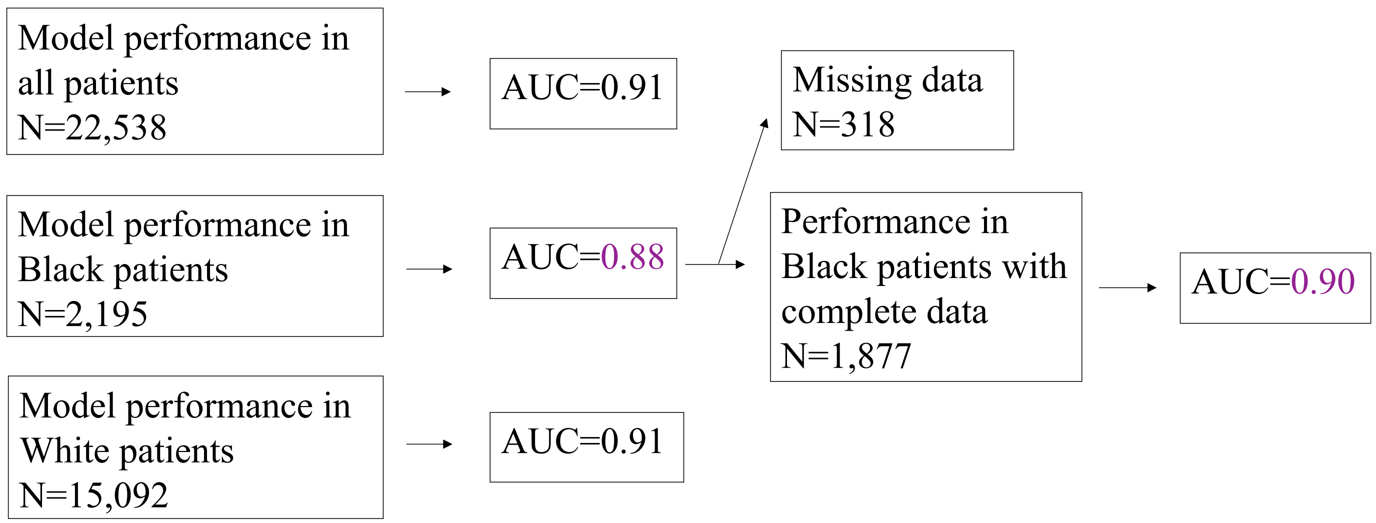

**Examples of individual patient predictions**

The models can also provide an individual prediction for a given patient at every time point. Using Shapley values, the features contributing to the prediction can be highlighted (appendix p 29). For example, one of the patients who subsequently developed preeclampsia had systolic and diastolic blood pressure, uric acid, and chronic hypertension as the most significant factors contributing to the model's predictions. In another example, a patient who remained normotensive throughout pregnancy had systolic and diastolic blood pressure and uric acid as the most important features contributing to the prediction.

Fig. S2. Examples of individual patient predictions A. Patient who developed preeclampsia B. Patient who was normotensive.

A. True positive B. True negative

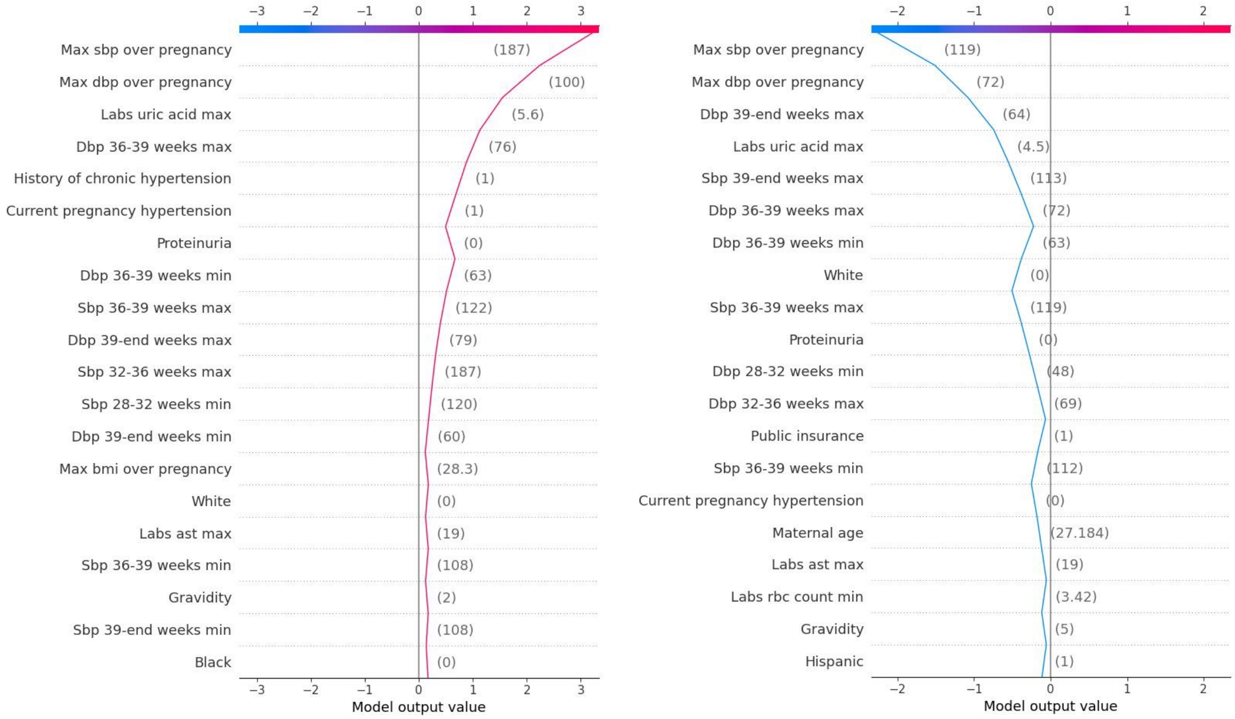

**Fig. S3.** Shapley values can be plotted by individual feature to show how the value of that feature affected the final model output. Every point is an individual delivery, and higher SHAP values show an increased effect toward a positive preeclampsia prediction. A variation on the y-axis at the same point on the x-axis reflects feature interaction. **A.** Relationship between the maximal systolic blood pressure during pregnancy, proteinuria, and risk for preeclampsia **B.** Relationship between maternal age, parity, and risk for preeclampsia. **C.** Relationship between maternal age, gravidity, and risk for preeclampsia. **D.** Relationship between maternal age, interpregnancy interval, and risk for preeclampsia. **E.** Relationship between RBC count during pregnancy and risk for preeclampsia

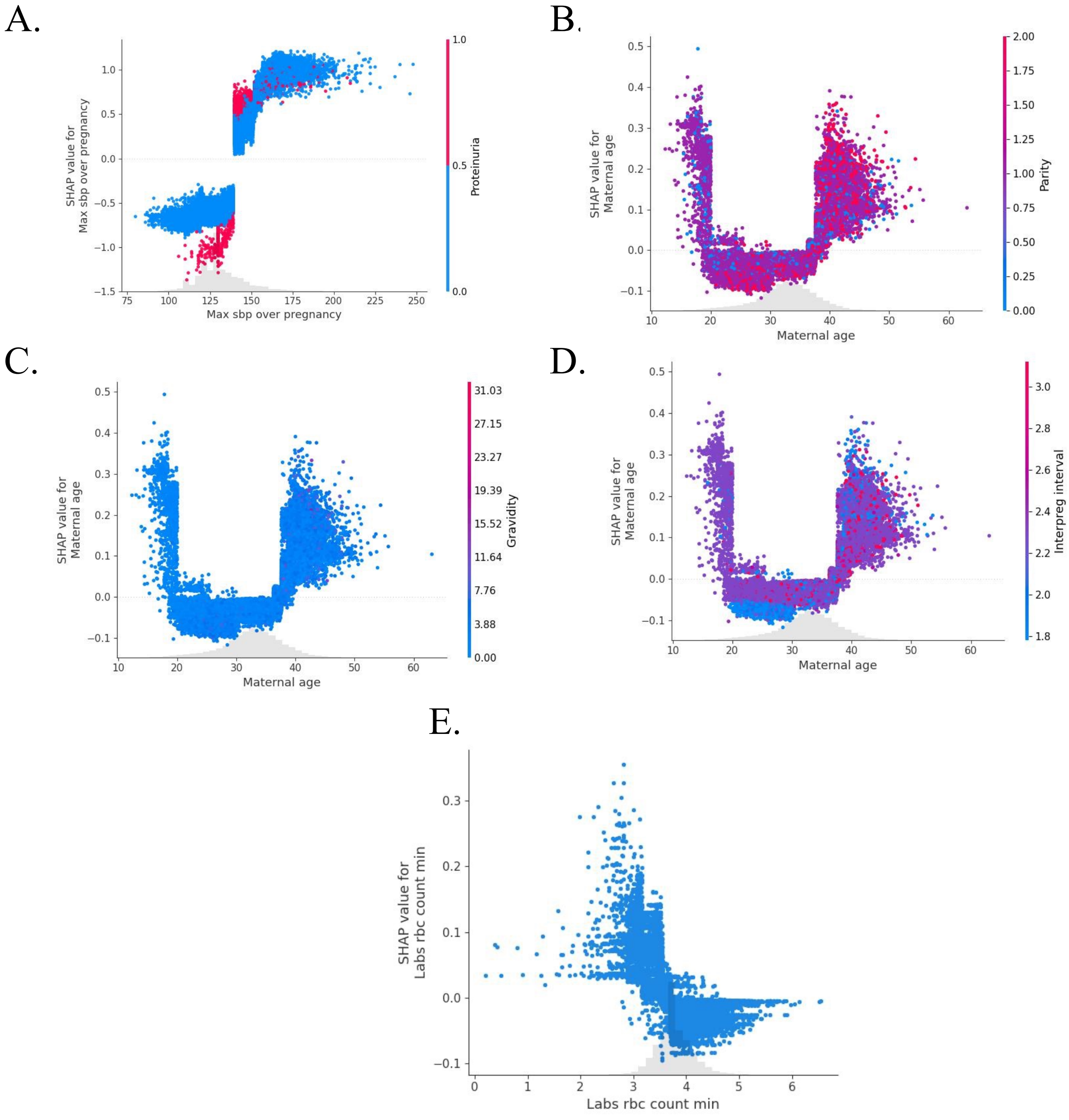

Supplementary Discussion

In light of AI bias and the selective disadvantage of individuals with self-reported Black race, multiple strategies have been developed to ameliorate the perpetuation of racial bias, including removing race as a variable. One study designed a novel method to account for the disproportionately higher risk of preeclampsia in Black patients; however, a major limitation of that work is the lack of data about medications, laboratory results, height, and weight of the patients.[3] We investigated the role of race in combination with other sociodemographic variables and employed best practices to develop highly performant and equitable tools. Black individuals are at higher risk for preeclampsia-related morbidity and mortality compared to White and Asian individuals.[4,5] Thus, we evaluated the predictive power of our models when using sociodemographic variables like type of insurance, quality of prenatal care, and substance use. The fact that the predictive power of the algorithm slightly increased with the addition of race suggests that Black individuals may be at higher risk for preeclampsia due to factors other than the sociodemographic and clinical variables we used. Another study has also demonstrated that removing race from machine learning models for preeclampsia risk prediction decreases the accuracy of the predictions.[6] Other prior work has shown that the elevated preeclampsia risk remains in Black individuals with higher income and socioeconomic status, suggesting the role of stress, innate biologic factors (e.g., genetic predisposition), and additional factors associated with systemic racism. [7] In addition, Black patients tend to have a higher proportion of missing data which may make research findings challenging to interpret.[3] We also find higher missingness in Black patients compared to the rest, which could be related to receiving care outside of the system or presenting later in pregnancy. When investigating the predictive power of the models, we found that they are highly performant in Black patients with lower levels of data missingness, suggesting that access to care and improving data availability in this patient population will be important strategies for the successful and equitable implementation of our models.

External validation is a major concern in modeling studies,[8] and most published models have lower predictive power in external validation due to changes in the feature definitions and overfitting of the model in the original population. [8] Thus, to mitigate the risk of overfitting, we selected AUC as a measure of the performance of the models and monitored the effects of techniques such as regularization and cross-validation, which reduce overfitting. In addition, as the models were developed using structured EHR data, we maintained uniform variable definitions and data processing strategies when we tested our tool on data from two independent community hospitals. This strategy has also demonstrated reproducible results.[9] The slight decrease in performance metrics in Cohort 2 as compared to Cohort 1 is due to a larger proportion of missing data in Cohort 2, potentially due to some outpatient offices using paper medical records.

Supplementary References

1. Akiba T SS, Yanase T, Ohta T, Koyama M. Optuna: A Next-generation Hyperparameter Optimization Framework. *arXiv* 2019; **1907.10902**.
2. He K, Zhang X, Ren S, Sun J. Deep Residual Learning for Image Recognition. *arXiv* 2015; **1512.03385**.
3. Bennett R, Mulla ZD, Parikh P, Hauspurg A, Razzaghi T. An imbalance-aware deep neural network for early prediction of preeclampsia. *PLoS One* 2022; **17**(4): e0266042.
4. Wright D, Syngelaki A, Akolekar R, Poon LC, Nicolaides KH. Competing risks model in screening for preeclampsia by maternal characteristics and medical history. *Am J Obstet Gynecol* 2015; **213**(1): 62 e1- e10.
5. Sandstrom A, Snowden JM, Bottai M, Stephansson O, Wikstrom AK. Routinely collected antenatal data for longitudinal prediction of preeclampsia in nulliparous women: a population-based study. *Sci Rep* 2021; **11**(1): 17973.
6. Ansbacher-Feldman Z, Syngelaki A, Meiri H, Cirkin R, Nicolaides KH, Louzoun Y. Machine-learning-based prediction of pre-eclampsia using first-trimester maternal characteristics and biomarkers. *Ultrasound Obstet Gynecol* 2022; **60**(6): 739-45.
7. Fasanya HO, Hsiao CJ, Armstrong-Sylvester KR, Beal SG. A Critical Review on the Use of Race in Understanding Racial Disparities in Preeclampsia. *J Appl Lab Med* 2021; **6**(1): 247-56.
8. Snell KIE, Allotey J, Smuk M, et al. External validation of prognostic models predicting pre-eclampsia: individual participant data meta-analysis. *BMC Med* 2020; **18**(1): 302.
9. Li S, Wang Z, Vieira LA, et al. Improving preeclampsia risk prediction by modeling pregnancy trajectories from routinely collected electronic medical record data. *NPJ Digit Med* 2022; **5**(1): 68.
